# Providing an e-cigarette starter kit for smoking cessation and reduction as adjunct to usual care to smokers with a mental health condition: Findings from the ESCAPE feasibility study

**DOI:** 10.1101/2024.09.13.24313612

**Authors:** Dimitra Kale, Emma Beard, Anna-Marie Marshall, Jodi Pervin, Qi Wu, Elena Ratschen, Lion Shahab

## Abstract

**Background:** Smoking rates in the UK have declined steadily over the past decades, masking considerable inequalities, as little change has been observed among people with a mental health condition. This trial sought to assess the feasibility and acceptability of supplying an electronic cigarette (e-cigarette) starter kit for smoking cessation as an adjunct to usual care for smoking cessation, to smokers with a mental health condition treated in the community, to inform a future effectiveness trial.

**Methods:** This randomised controlled feasibility trial, conducted March-December 2022, compared the intervention (e-cigarette starter kit with a corresponding information leaflet and demonstration with Very Brief Advice) with a ‘usual care’ control at 1-month follow-up. Participants were ≥18 years, receiving treatment for any mental health condition in primary or secondary care in three Mental Health Trusts in Yorkshire and one in London, UK. They were also willing to address their smoking through either cessation or reduction of cigarette consumption. The agreed primary outcome measure was feasibility (consent∼15% of eligible participants; attrition rate<30%). Acceptability, validated sustained abstinence and ≥50% cigarette consumption reduction at 1-month, were also evaluated and qualitative interviews conducted to further explore acceptability in this population.

**Results:** Feasibility targets were partially met; of 201 eligible participants, 43 (mean age = 45.2, SD = 12.7; 39.5% female) were recruited (21.4%) and randomised (intervention:48.8%, n=21; control:51.2%, n=22). Attrition rate was 37.2% at 1-month follow-up and was higher (45.5%) in the control group. At follow-up (n=27), 93.3% (n=14) in the intervention group and 25.0% (n=3) in the control group reported e-cigarette use. The intervention was well received with minimal negative effects. In intention-to-treat analysis, validated sustained abstinence at 1-month was 2/21 (9.5%) and 0/22 (0%) and at least 50% reduction in cigarette consumption 13/21 (61.9%) and 3/22 (13.6%), for the intervention and control group, respectively. Qualitative analysis of participant interviews (N=5) showed the intervention was broadly acceptable, but they also highlighted areas of improvements for the intervention and trial delivery.

**Conclusions:** Offering an e-cigarette starter kit to smokers with a mental health condition treated in the community was acceptable and largely feasible, with harm reduction outcomes (i.e. switching from cigarette smoking to e-cigarette use and substantial reduction in cigarette consumption) favouring the intervention. The findings of the study will be used to help inform the design of a main trial.

**Trial Registration:** Registry: ISRCTN

Registration number: ISRCTN17691451

Date of registration: 30/09/2021

## Background

Despite smoking prevalence in the UK general population declining steadily over the past five decades, now standing at ∼14% (1), little change has been observed among people with a mental health condition (2, 3), who are still more than twice as likely to be smokers as the general population (4). Combined with high levels of nicotine dependence (5), which result in generally high cigarette consumption, this leads to substantially increased risks of smoking-related morbidity and premature mortality in people with a mental health condition (2). Although people with a mental health condition are similarly motivated (6) and able (7) to quit smoking as those without a mental health condition, quitting smoking can be difficult due to limited access to support and high dependence. Thus, there is a clear need to develop better and more tailored support strategies to aid smoking cessation in this population. Electronic cigarettes (e-cigarettes), which deliver nicotine without most of the harmful substances in tobacco smoke, have been recognised as a potentially helpful tool for smoking cessation (8).

E-cigarette use (vaping) is increasingly being recorded by stop smoking services in the UK (9), and may be more appealing to people with a mental health condition, who are more likely to have tried vaping and be current users than smokers in the general population (10). Potential reasons for this include that e-cigarettes are relatively cheap compared with cigarettes and other cessation treatment (11) and that they offer a simple stand-alone treatment that is intuitive to use. Furthermore, since e-cigarettes simulate the sensory input from cigarettes and allow users to control the dose (unlike most nicotine replacement therapies (NRTs)), they may appeal to more dependent smokers who have hitherto struggled to quit with existing cessation treatments (12). Thus, e-cigarettes may offer a potential solution to reduce smoking and encourage cessation in mental health care settings by functioning as a safer alternative to cigarettes (13).

In the general population, accumulating evidence suggests that e-cigarettes are as effective as, or even more effective than, NRT in aiding smoking cessation, both from randomised controlled trials (RCTs) (14) and real-world studies (15). In addition, there are small observational studies, which support their use for people with a mental health condition. Work carried out in Italy (16), the US (17) and Australia (18) found that e-cigarettes are potentially effective for smoking cessation and reduction among smokers with a serious mental health condition. More recently, a UK pilot study investigating the utility of e-cigarettes as a harm reduction intervention in people with psychotic disorders reported a significant reduction in average number of cigarettes smoked per day between baseline and 6-week follow-up, supporting the notion of e-cigarettes as a useful harm reduction tool for this population (19). These studies did not find any evidence that e-cigarettes had adverse effects on mental health, further underlining their utility as a safe smoking cessation aid for smokers with a mental health condition. However, while these preliminary results are encouraging, to date no adequately powered RCT has assessed the effectiveness of e-cigarettes as a long-term harm reduction and smoking cessation tool for people with a mental health condition.

The overall aim of this research was to undertake a feasibility study to evaluate the feasibility and acceptability of supplying e-cigarette starter kits, along with brief verbal and written advice on e-cigarette use, as an adjunct to usual care for smoking cessation in smokers with a mental health condition treated in the community prior to undertaking a full RCT (E-cigarettes for Smoking Cessation And reduction in People with mEntal illness (ESCAPE trial)). The following research objectives were specified:

1. Assess the feasibility of conducting a full RCT by estimating recruiting (eligible patients who were invited to take part in the study), consenting (those who consented to participate), attrition rates as well as treatment adherence in the intervention group, and contamination in the control group.
2. Investigate the acceptability of trial procedures and the intervention in terms of written materials, verbal content and e-cigarette provided.
3. Explore signals pertaining to the potential efficacy of the intervention.

## Methods

### Study design

This was a feasibility study using an RCT design, comparing the intervention (an e-cigarette starter kit, brief demonstration, verbal and written information on e-cigarette use as an adjunct to usual care) and control (usual care) at 1-month follow-up. Participants were recruited between March and December 2022. The recruitment stopped at December 2022 due to funding constraints and planned timelines to enable review and progression to the full trial. Additionally, we conducted qualitative interviews to explore the experience of service users and researchers delivering the intervention to refine the intervention accordingly. Ethical approval was granted by the NHS HRA (REC ref:21/NE/0202).

### Participants

Trust researchers identified potential participants via health records prior to attendance at their annual care programme approach review or physical health screening appointment. Potential participants were then sent participant information sheets about the study by their trust one week prior to their appointment. Before their appointment, a trust researcher approached them to assess their interest in the study and eligibility. As interest and motivation to participate in smoking cessation studies can fluctuate (20), those who declined on the first occasion were offered participation in the study up to a total of three times, through a letter, follow-up call, and, if possible, a text message. Following the eligibility check, the trust researcher briefly explained the trial and consented participants. Only eligible, consented participants were then asked to complete a brief baseline questionnaire and randomisation occurred after completion of baseline questionnaire. The baseline questionnaire could be completed either on paper or online via REDCap links.

### Inclusion/exclusion criteria

Participant inclusion criteria were: i) aged 18 and over, ii) self-reported current (in the past 7 days) cigarette smoking, iii) a diagnosis of a mental health condition and currently receiving treatment for this in primary or secondary care (community mental health teams) validated by their health care records and, iv) a willingness to address their smoking behaviour either by attempting to quit, or by reducing their cigarette consumption. Participants were excluded if they: i) had an inpatient admission in the last three months, ii) self-reported current regular (at least weekly) use of e-cigarette, iii) self-reported participation in other smoking cessation study, iv) were receiving current treatment for comorbid drug or alcohol problems, v) had a diagnosis of Alzheimer’s disease or dementia, and vi) were pregnant or breastfeeding.

### Setting

Participants were recruited from three Mental Health Trusts in Yorkshire and one in London, UK.

### Randomisation

The intervention allocation was determined by computer block-randomisation to ensure that each trial site had an equal proportion of intervention and control group participants. Randomisation occurred after consent to take part in the study had been obtained via opening of consecutively numbered opaque envelopes containing information about allocation. The allocation slip was also double folded, and envelope sealed with a signature on the envelope sealed flap. Allocation was concealed until after completion of the baseline questionnaire. Researchers informed the participant and the clinical team of allocation.

### Blinding

Participants and researchers and clinical staff administering the intervention could not be blinded due to the nature of the intervention and study design. As follow-up questionnaires differed for intervention and control groups, outcome assessment was only blinded to researchers for questionnaires self-completed online rather than over the phone. However, the study’s statistician was blinded to participants’ allocation.

### Control group

Control group participants received usual care from their clinician. While we did not explicitly assess what this entailed (given that we would not have had power to analyse specific differences), as per u NICE guideline NG209 (21), at a minimum standard this would involve evidence-based Very Brief Advice to stop smoking, comprising the three As (Ask and record smoking status; Advice on the best way of quitting and; Act on patient response to build confidence (22, 23)) and referral to in-house or external specialist stop smoking services. In-house and external specialist stop smoking services may offer more tailored behavioural support for smoking cessation, including advice and information on smoking cessation aids such as NRT. However, we did not collect data on how many participants were referred to stop smoking services or on the specific details of the usual care offered by the participating trusts in the feasibility trial. As part of the intervention, all participants, irrespective of their motivation to stop, were encouraged by the trust researcher to consider quitting and to set up a target quit date within a week after randomisation, and those who did not wish to set a target quit date were encouraged to reduce cigarette consumption.

### Intervention group

Intervention group participants were offered an e-cigarette starter kit comprising of a third-generation e-cigarette (Aspire PocketX) with a four-week supply of a choice of: a) nicotine strength e-liquid (<u>3 options</u>: 6mg/ml, 10mg/ml, 18mg/ml) and b) flavours (<u>3 options</u>: tobacco, fruit, menthol) as an adjunct to usual care. They also received a verbal explanation and demonstration, along with an information leaflet on how to use the e-cigarette. This was delivered by a clinical member of staff in the context of a pre-existing clinical appointment. The information leaflet included details on what an e-cigarette is, what to expect from it, how to set it up, and how to use it correctly (a copy of the information leaflet is provided in the Supplementary files). All participants, irrespective of their motivation to stop, were encouraged to consider quitting and to set a target quit date within a week after randomisation. Participants were asked to start using the e-cigarettes as soon as possible and to seek out local or online vape shops to obtain further e-liquid, suited to their individual needs and flavour preference. Participants who did not wish to set a target quit date were encouraged to use the e-cigarette to reduce cigarette consumption as soon as possible.

### Data collection

Questionnaires at baseline and follow-up were initially administered by researchers either online (data were captured and managed by the REDCap electronic data system) (24, 25) or in person at the site using a paper-based version. The options of completion via telephone and home visit were added in July 2022.

### Follow-up

At 1-month after randomisation participants were asked to complete another brief questionnaire, either online or via telephone. Participants were followed up with up to three reminders to complete the 1-month follow-up.

To support engagement, all participants (both in intervention and control group) received a £10 love2shop voucher for completing the baseline assessment and a £10 love2shop voucher for completing the 1-month follow-up assessment.

### Measures

#### Baseline measures

Mental health diagnosis obtained from health care records. All the other measures were self-reported. Sociodemographic characteristics included age, sex, ethnicity, employment status and education attainment.

Smoking-related characteristics included nicotine dependence measured by the Strength of Urges to Smoke Scale (26) and number of cigarettes smoked per day, motivation to quit measured by the Motivation to Stop Scale (27), age started smoking, smoking duration, past year quit attempts and ever vaping as smoking cessation aid. Mental health condition symptoms were assessed using the Patient Health Questionnaire (PHQ-9) (28) and Generalised Anxiety Disorder (GAD-7) scale (29).

### Outcome measures

#### Feasibility

To assess recruitment and consenting rates, we recorded the number of eligible people who were invited, and the number who consented to take part. We sought to consent a minimum of ∼15% of eligible participants. As this is a harder to reach population, this is slightly lower than the consenting rate typical in clinical trials in general patient populations in the UK (30). We also assessed recruitment rates at each Trust. We sought to recruit around six participants per Trust per month to ensure a reasonable timeframe for delivery of a full RCT. Attrition rate was measured by recording the proportion of participants who fail to complete the 1-month follow-up assessment. To achieve a sufficiently robust effect estimate, which can be affected by high attrition rates in intention-to-treat analyses (31), we sought an attrition rate below 30%, which is typical for smoking cessation trials in this population (32). Finally, adherence to treatment and contamination was measured by recording the proportion of participants who used/were using an e-cigarette at 1-month follow-up in the intervention and control groups, respectively.

#### Acceptability of trial procedures and the intervention

To assess acceptability of the trial procedures and intervention, qualitative interviews were conducted with a sample of five participants and five researchers delivering the intervention. An interview protocol (Supplementary Table 1) was designed to gain insights into participants’ and researchers’ experience with the trial, the intervention, and barriers and facilitators of success, both in terms of trial procedures and the intervention content. All participants at baseline were asked if they agree to be interviewed after the 1-month follow-up. Of those who agreed, five attended an interview; two from the control group and three from the intervention group (three were females and two males). Similarly, all researchers were asked if they agreed to be interviewed after the 1-month follow-up, and five mental health nurses (all female) from two trusts attended an interview. All interviews were conducted over the telephone by three researchers from the University of York, lasted between 20 and 40 minutes, were audio-recorded and transcribed verbatim. Interviewees received £10 for participating in the interviews.

Further, among participants in the intervention group, acceptability of the intervention in terms of written materials, verbal content and e-cigarette provided was measured at 1- month follow-up with a questionnaire based on other acceptability-related research conducted in this population ((33); 14-items (Table 2), answer options 5-point Likert-scale from 1=not at all to 5=extremely).

**Table 1:** Characteristics of participants overall and as function of group at baseline.

| Characteristic | Overall (n=43) | Control group (n=22) | Intervention group (n=21) |
| --- | --- | --- | --- |
| **Sociodemographic** |  |  |  |
| Age, M (SD) | 45.2 (12.7) | 42.9 (12.6) | 47.7 (12.6) |
| Female sex, % (n) | 39.5 (17) | 36.1 (8) | 42.9 (9) |
| White ethnicity, % (n) | 76.7 (33) | 72.7 (16) | 81.0 (17) |
| Post-16 educational qualifications, % (n) | 16.3 (7) | 13.6 (3) | 19.0 (4) |
| Employed, % (n) | 27.9 (12) | 45.5 (10) | 9.5 (2) |
| **General and Mental Health characteristics** |  |  |  |
| PHQ-9, M (SD) | 12.3 (6.7) | 10.5 (6.7) | 14.2 (7.5) |
| GAD-7, M (SD) | 9.2 (6.2) | 7.9 (5.2) | 10.6 (7.1) |
| **Smoking-related characteristics** |  |  |  |
| Years smoked, M (SD) | 23.7 (13.6) | 23.5 (11.9) | 24.0 (15.5) |
| Age started smoking, M (SD) | 18.0 (9.0) | 15.4 (3.6) | 20.6 (11.9) |
| Strength of urges to smoke, M (SD) | 3.4 (1.0) | 3.4 (1.0) | 3.5 (1.1) |
| Time spent with urges, M (SD) | 3.4 (1.4) | 3.7 (1.3) | 3.1 (1.5) |
| Cigarettes smoked per day, M (SD) | 19.6 (10.9) | 20.0 (12.8) | 19.1 (8.9) |
| Motivation to stop smoking, M (SD) | 5.2 (1.7) | 4.8 (1.6) | 5.6 (1.7) |
| Past year quit attempt, % (n) | 37.2 (16) | 40.9 (9) | 33.3 (7) |
| Past year e-cig use for smoking cessation, % (n) | 27.9 (12) | 27.3 (6) | 28.6 (6) |

Motivation to stop was measured with the motivation to stop scale (scores 1-7) with higher score presenting higher motivation to quit cigarette smoking.
Strength of urges to smoke (scores 0-6) with higher score presenting higher strength of urges to smoke.
Time spent with urges (scores 0-6) with higher score presenting more time spent with urges.
PHQ-9: scores represent: 0-5 mild, 6-10 moderate, 11-15 moderately severe, >15 severe depressive symptoms.
GAD-7: Scores represent: 0-5 mild, 6-10 moderate, 11-15 moderately severe, 15-21 severe anxiety.

**Table 2:**
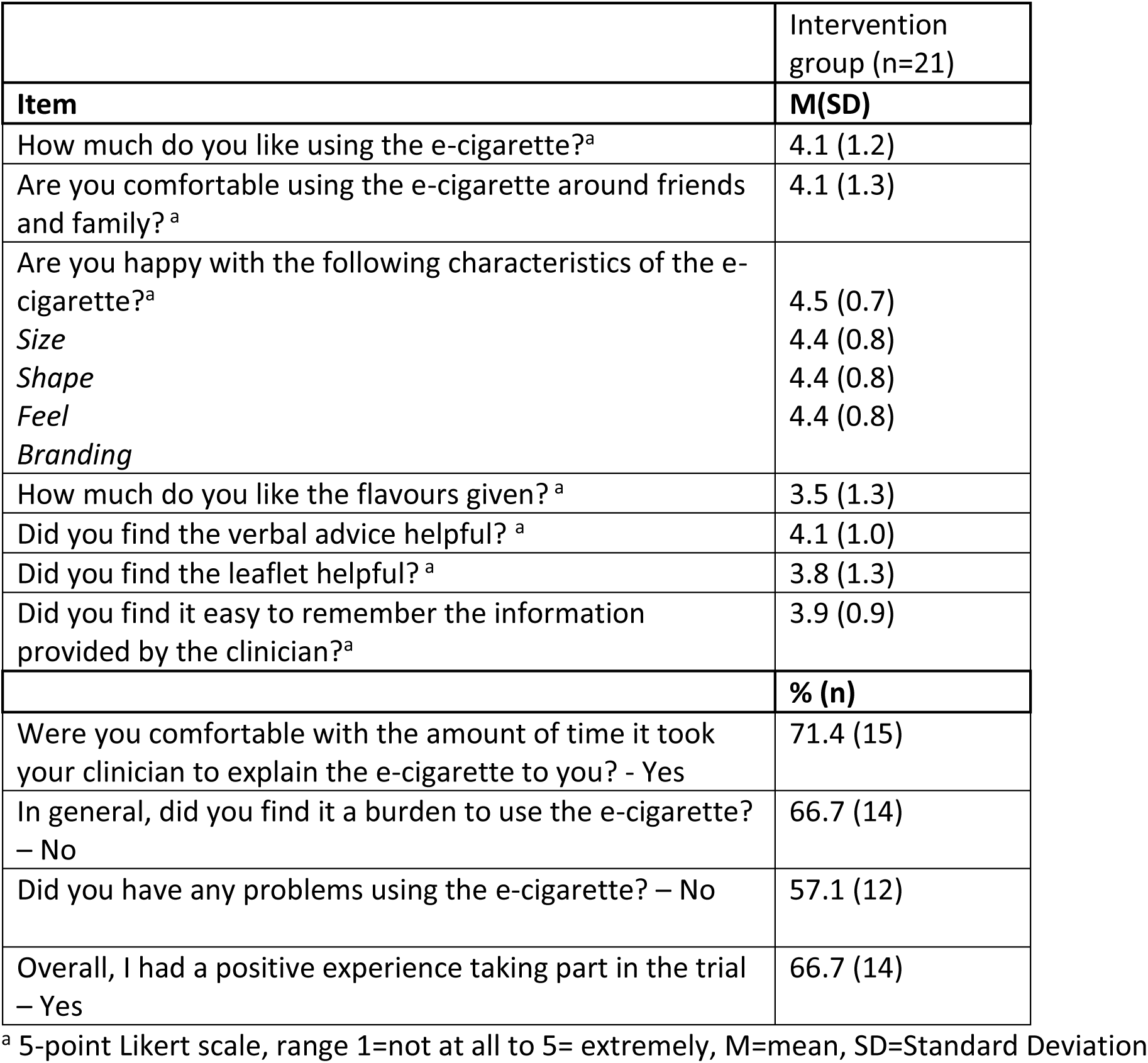
Acceptability of the intervention among participants in the intervention group.

#### Changes in mental health symptoms and experience of adverse events

Additionally, we assessed changes in participants’ mental health symptoms (PHQ-9 (28) and GAD-7 (29)), general mood and physical symptoms (MPSS, (34, 35)), and experience of adverse events based on previous vaping trials (12).

#### Potential efficacy

To explore signals of potential intervention efficacy at 1-month follow-up, we recorded: i) CO validated (<10ppm) sustained abstinence for weeks 2-4 from enrolment date or target quit date (for those who set a date within one week of enrolment), equivalent to the standard measure in UK stop smoking services (36); ii) point prevalence (24h) abstinence; and iii) the proportion achieving 50% smoking reduction, a common outcome measure used in vaping cessation studies (37). At the 1-month follow-up, we also assessed how many participants had set a quit date in the past month and how many had used NRT during that time.

#### Health economics

To identify appropriate instruments and assess the feasibility of collecting health economic data, we collected resource use data for delivering the intervention and usual care, using both trial records and a bespoke service use questionnaire. The EQ-5D-5L questionnaire was administered to collect data that enable the estimation of the quality-adjusted life year, which is the most commonly used health outcome measure in economic evaluations (38, 39).

### Sample size

The target sample size for this randomised controlled feasibility trial was 72, with 36 participants allocated to each group. In a full RCT, we would assume an effect size of OR (odds ratio) of 1.9 (pooled estimate based on e-cigarette vs placebo e-cigarette trials: (16, 40) for the outcome of 1-month continuous abstinence rate. This would result in an absolute predicted risk difference of 8.2%, assuming a 1-month abstinence rate in the control group of 11.4% (based on EAGLES trial (41)) and 19.6% in the intervention group. The feasibility sample size would be sufficient to produce a one-sided confidence interval that excludes an 8.0% difference in the event of a zero effect of the intervention on abstinence at 1 month, assuming 11.0% reported abstinence in each of the two groups. The estimate obtained in the feasibility trial is not used to directly estimate the intervention effect but to determine whether proceeding to a trial is worthwhile, based on the one-sided confidence interval approach (42). Because the target sample size was not achieved, this analytic approach could not be followed. As recommended by our trial statistician based on relevant literature (43), several two-sided confidence intervals of different precision were therefore calculated instead.

### Analyses

Baseline characteristics and relevant follow-up measures (adverse events, mood and physical symptoms) are summarised using percentages and frequencies for categorical variables and means and standard deviations for continuous variables and the groups (intervention vs control). Differences between groups were compared using t-tests, Mann Whitney U tests, or Fisher’s Exact test as appropriate. Alpha was set to 0.05.

To assess feasibility, we present frequency and percentages for the number of eligible smokers who i) were invited to take part and consented/completed the baseline assessment; ii) were recruited at each trust per month; iii) attended and completed the 1- month follow-up; iv) had used or were using an e-cigarette at 1-month follow-up. The feasibility criteria as cited earlier were i) consent ∼15% of eligible participants; ii) recruit ∼6 participants/Trust/month; iii) attrition rate <30%.

To assess acceptability of the trial procedures and intervention, interview data were analysed using a deductive Thematic Analysis (44) approach, and the analysis was informed by the topic guide. Additionally, acceptability of the intervention among participants in the intervention group is summarised using frequencies and percentages.

Differences between groups in sustained and point prevalence smoking abstinence at 1- month follow-up was assessed using the one-sided confidence interval approach (42). This analysis was based on the assumption that the sample identified in the sample size calculation, n=72, were recruited. One-sided 80% confidence intervals were derived. If the upper confidence interval excludes 8% (the clinically significant effect in the sample size calculation) difference, then it could be concluded that the difference between the two groups was 0. However, as this study did not reach the sample size of n=72, this planned confidence interval approach could be flawed as the study was underpowered. Thus, in an unplanned analysis, several two-sided confidence intervals of different lengths were calculated to determine at which level a treatment effect might be present (43).

Generalised linear models (with intervention allocation specified as between-group factor and time as within-group factor) was used to determine reductions in cigarette consumption and changes in mental health measures from baseline to 1-month follow-up. Analyses of smoking-related outcomes followed the intention-to-treat principle with treating those lost to follow-up as smokers/not having changed their consumption. Additionally, we also undertook a complete case analysis. Data were analysed in SPSS 28.0

To estimate the cost of the intervention and usual care, the quantity of each type of healthcare resource used during the trial period was valued by attaching a corresponding unit cost obtained from either the ESCAPE trial or established national sources (45-48). The total costs were summed and divided by the number of participants to calculate the mean cost for each group. Costs were expressed in British pounds (£) at 2021/2022 prices.

## Results

### Baseline participants’ characteristics

All participants had a diagnosed mental health condition and were receiving treatment in primary or secondary care, as indicated by their health records. However, we have exact diagnoses for only some participants: 13 with depression, five with bipolar disorder, four with psychosis, seven with schizophrenia, one with persistent delusional disorder, and one with personality disorder with antisocial traits. We were unable to retrieve the exact mental health diagnoses of the remaining participants. We did not obtain any information regarding the specific treatments they were receiving. Table 1 shows participants’ characteristics overall and as a function of group assignment. The average age of participants was 45.2 (Standard Deviation (SD)=12.7), the majority were male (60.5%) and of white ethnicity (76.7%). Post-16 educational qualifications were held by 16.3%, and 27.9% were employed. In terms of general and mental health, the average PHQ-9 and GAD-7 scores was 12.3 (SD=6.7) and 9.2 (SD=6.2) respectively, suggesting moderate depression and anxiety. For smoking-related characteristics, participants had smoked for an average of 23.7 years (SD=13.6), starting at an average age of 18.0 years (SD=9.0). They smoked an average of 19.6 cigarettes per day (SD=10.9), and their mean motivation to stop smoking was 5.2 (SD=1.7). Regarding past year smoking cessation efforts, 37.2% had attempted to quit, with slightly more in the control group (40.9%) than in the intervention group (33.3%). Additionally, 27.9% had used e-cigarettes for smoking cessation in the past year, with similar proportions in both groups Additionally, 37.2% had attempted to quit and 27.9% had used e-cigarettes for smoking cessation in the past year.

### Feasibility

#### Recruitment and consenting

Two hundred and one smokers with a mental health condition treated in the community were eligible and invited to participate. Of these 43 (21.4%) consented, completed baseline assessment, and were randomised (21 to the intervention group and 22 to the control group; figure 1). The overall monthly recruitment rate was below expectation (six participants/Trust/month) at 1.95 participants per month per Trust; however, this rate differed greatly between sites, from three participant per month at one of the Yorkshire sites to only 0.5 participants per month at the London site.

**Figure 1.**
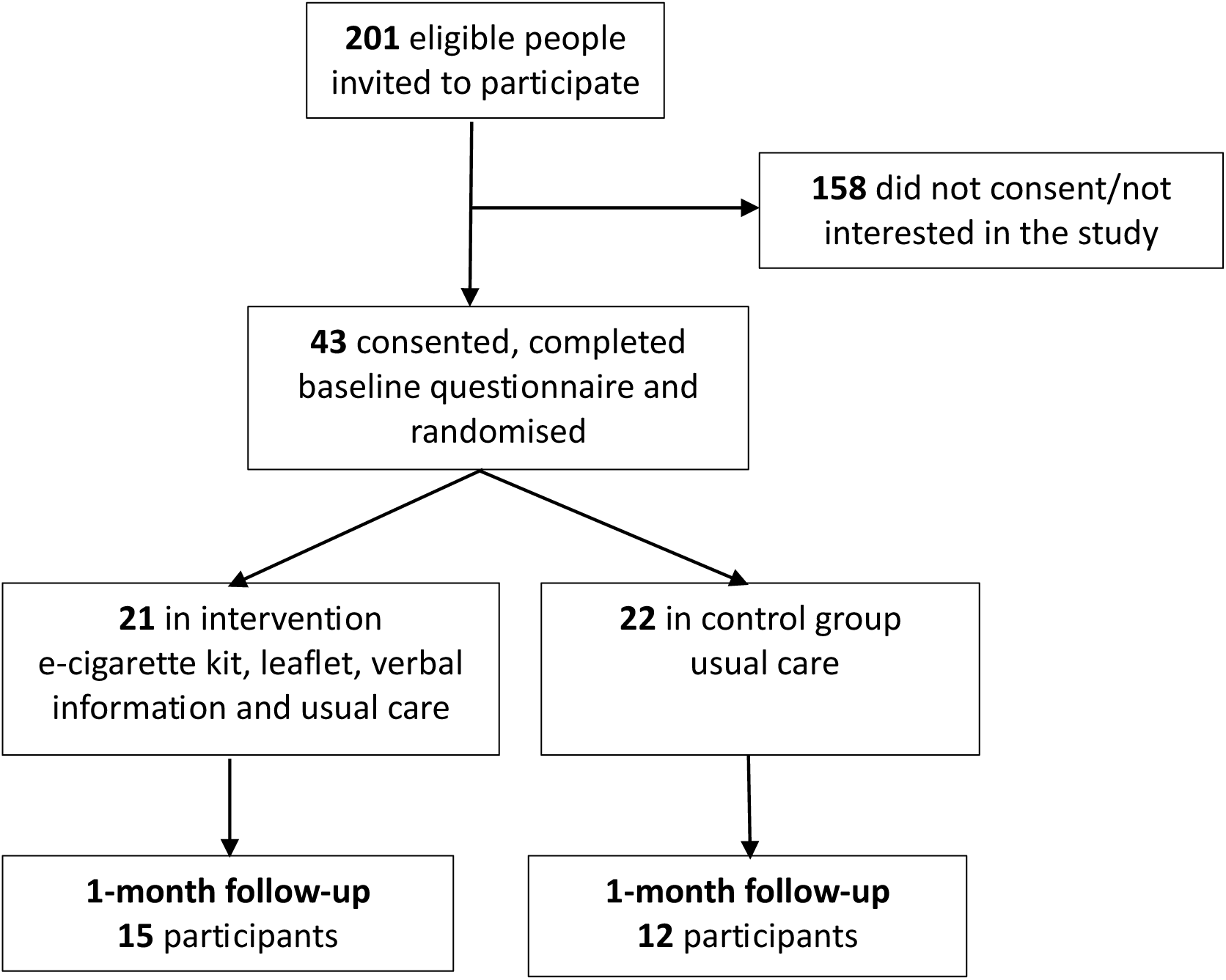
CONSORT flowchart of participant recruitment.

#### Attrition

Overall attrition rate was 37.2% (n=16) at 1-month follow-up, and attrition was higher in the control (45.5%; n=10) than intervention (28.6%; n=6) group.

#### Adherence to treatment and contamination

Of participants followed-up at 1-month (n=27), 14 (93.3%) in the intervention group reported using the e-cigarette in the last month, while three (25.0%) participants of the control group also reported e-cigarette use at follow-up.

### Acceptability of trial procedures and the intervention

#### Qualitative assessment of intervention and trial procedures acceptability

Four main themes depicted in Table 3 were derived, with further details provided below.

**Table 3.** Qualitative interviews: Themes and related codes of qualitative analysis.

| Theme | Related codes | Related quotes |
| --- | --- | --- |
| 1. Improvements to the trial's materials and procedure | Unhappy with questionnaire content and length | P5(I): Felt uncomfortable completing the questionnaires.<br>P5(I): Felt uncomfortable answering question about mental health.<br>P5(I): Questionnaire long and questions repetitive.<br>P5(I): Did not understand why we asked questions unrelated to smoking - should explain better.<br>P3(I): Didn't expect so many questions.<br>P3(I): Surprised by questionnaire content and length.<br>P4(I): Would like more time to complete the questionnaire.<br>R2: I found that some of the questions on the on the follow up. I've kind of tend to repeat. Being a bit repetitive. They definitely are repetitive. They've had a few comments. That kind of oh, haven't you already asked me this? |
|  | Researcher's support helpful during questionnaire completion | P2(C): I liked that researcher helped me complete the questionnaire.<br>P3(I): Help to complete the questionnaire is important due to the length.<br>R1: There was a preference for in person data collection... there were some issues with the content or the questionnaires that they struggled with. They felt it was repetitive and lengthy.<br>R3: A lot of people what they've wanted to do it online. However, did explain that in secondary care in person assistance was usually required.<br>R2: In reality they end up being done with a researcher because it's too much. It's too much for them to do independently. |
|  | Views of interactions with clinical staff | P1(C): Need to be clearer with patients about what we are offering from the start of communication.<br>P1(C): Mental Health teams told patients this was smoking cessation support rather than research.<br>P1(C): CPN relayed ESCAPE incorrectly as support rather than research. |
|  | Initial contact preferences – telephone | P3(I): Did not read letter.<br>P3(I): Phone call from researcher fundamental in decision to take part.<br>P3(I): Requires spoken explanation - preferred over online or post "sinks in better".<br>R3: Some people are not very tech savvy and would rather have the option of reading to it themselves and make it a decision. Other people like to speak to people, I think at the moment people the majority would perhaps appreciate speaking to somebody.<br>R5: The letters definitely didn't have much of an impact. We didn't get anyone replying back to them. |
| 2. Evidence of intervention acceptability | Happy with clinician appointment | P3(I): Yeah, I mean they were there to support me.<br>P3(I): Yeah, she was because like she explained to me how to use it and I explained like using something like this seemed to help.<br>P4 (I): I think this support was good, the communication was good, and it was helpful. |
|  | Positive experience/feelings towards e-cigarette(s) | P3(I): It's been good it's been good it's given me more positivity...I haven't been craving I've not been craving as much.<br>P3(I): I'd say the benefits it's making me want to quit.<br>P3(I): Not only that cold weather's coming now and I'd rather sit in me house and smoke me vape.<br>P4(I): It was a good quality e-cigarette. I have had poor quality ones, but this was a good one.<br>P4(I): The e-cigarette is better for health. For safety wise it's good as well the e-cigarette.<br>P5(I): it's a good device.<br>R1: For the liquid and e-cig there seemed to be a good choice.<br>R1: The e-cigarette looked nice, it looks expensive, so I think they liked the look of it. |
|  | The leaflet was useful | P3(I): I did find it useful.<br>P4(I): No, I didn't use it because she explained to me how to use it and how to charge it and I have experience of my own which I had in the past so I know how to use it.<br>R1: It was really comprehensive. |
|  | Overall experience positive | P3(I): It's given me more positivity.<br>P3(I): You know it's given me encouragement.<br>P4(I): It was good, a good experience.<br>P5 (I): The process itself is a fantastic idea um I can't fault the fact of what's been done.<br>R3: All I know is that from. You know the people of going to study. They've been wanting to stop smoking, and I found it useful in one way, either way, you know, and I've had I I've had really positive feedback.<br>R2: From the follow up as well. and people have found it really useful. And what I found interesting is that actually it's almost like the idea. And the concept of using an e-cigarette and the fact that I guess I'm picking up on it's the fact that the health professional is giving someone any cigarette and saying that it's, you know it's a helpful and useful and safe thing to do. It's really powerful, because I think quite a lot of people just need that reassurance. |
| 3. Improvements to the intervention | Rejection and worry due to randomisation | P1(C): It is really interesting because as a result of my mental health issues and past traumas and things, because I didn't get the vape and the interaction it actually had the |
|  |  | opposite effect with me. I actually ended up smoking that week a lot more, it was like a sense of rejection.<br>P1(C): When they explained to me what it actually was I was on tenterhooks whether or not I was going to get this vape and then I got the appointment with Kim and I didn't get the vape and when I didn't I was so disappointed.<br>P1(C). The conversations I have had with practitioners and you have helped. Although with one of the practitioners, when I didn't get the vape I was disappointed that I was just going to get left again and for me I have been signed up with smoking cessation support but have never heard anything so it was like rejection all over again because I wasn't informed enough in the first instance I know it is a research project but when I first got referred I wasn't aware of that.<br>P3(I): The thing what was worrying me before that and they explained that what if she opened envelope and it wasn't so where do I go from there.<br>P5(I): In order for the if you weren't to get picked then I would imagine cause I'm not going to lie if I hadn't got picked I'd have prob- I'd have been on a right you know I'd have been really bad and sad.<br>R1: I think a couple were upset at follow-up. There was one that was really upset. I was quite surprised because I thought I had managed it well. I talk a lot about it and try to manage their expectations. They might feel that they don't want to say it at the time. I hope that we managed it as best we could, it's just one of those things. I think the voucher was a good idea and that helped and dividing it and giving half at baseline softened the blow. |
|  | More help using the e-cigarette required | P3(I): She gave me it we did the set up and things like that but she didn't actually turn it on... so when I got home and I tried to do things cause she had to check it three times and I got it didn't work it made me a little bit frustrated but after talking to (consultant's name) again I took it in and- and she got it up and running whereas I think if it happened again or she for somebody else I'd say set it up for her.<br>P3(I): If they go more into it with you and have you do a trial with different things and show you how to use the apparatus then I think it might have been better"<br>P5(I): Like I said I'm not able to get the effects of what this device is meant to do... I don't obviously want to get her in trouble or anything but no I wasn't sure I wasn't sure how to use the device.<br>R2: Doing real deep in inhalations, and then coughing and feeling a bit put off by that. And so, they were using it like |
|  |  | a cigarette. So, I don't know whether there was kind of a placing a little bit more emphasis on how you use it, and maybe how you set it up in the intervention, I think people would find that helpful.<br>R3: There are a lot of questions, and I think it would. I don't think. Is it the actual saying, talking about stopping, smoking can be done in 5 min, but the actual vape itself felt quite personal. So, I had a heck of a lot of questions. |
|  | On-going support/contact important | P1(C): For me it's not about the e-cigarette, it's about the support for stopping smoking because that's what's worked for me in the past.<br>P1(C): For me, the thing that I think will help is having that ongoing support... Just to add again with it being mental health services, we don't always absorb information. No disrespect, clinicians were fantastic but maybe we need reminders of the important stuff. I think a telephone call maybe 3-5 days after would be really good.<br>P4(I): Maybe she could have seen me again or catch-up with me to see how I was doing with my smoking or something. |
| 4. Issues with the e-cigarette | Side effects of e-cigarette | P3(I): It made back of my throat burn.<br>P3(I): Negative effects coughing. |
|  | Ongoing cost of e-cigarette is problematic | P2(C): Tobacco cheaper than e-cigarette.<br>P2(C): E-cigarettes are too expensive, can't afford ongoing cost.<br>P4(I): E-liquid expensive and would not be able to afford this or the e-cigarette without being given one for the trial. |
|  | Difficulty using the e-cigarette at home | P3(I): Couldn't work out how to use e-cigarette at home.<br>P3(I): Revisited clinician to understand how to use e-cigarette.<br>P4(I): Takes time to get used to the difference, e-cigarette very different to smoking, it is awkward when you start.<br>P5(I): Got liquid in mouth. |
|  | The e-cigarette is not very good | P5(I): E-cigarette isn't very good.<br>P5(I): Disappointed - faulty device.<br>P5(I): Would prefer a different device.<br>P5(I): Device not good compared to others on the market.<br>R2: People don't seem to like tobacco flavour. So, not keen on that. |

P=participant; I=intervention; C=control; R=researcher

#### 1. Improvements to trial’s materials and procedure

Three participants felt that the questionnaires were too long, and one would have liked more time to complete them. Researchers also reported that the questions were repetitive. Participants felt that researcher support was helpful during questionnaire completion. In-person and telephone contact were preferred over online engagement and letters, and researchers reported that in-person assistance was often required for questionnaire completion. Clinical staff outside of the team were reported to have relayed incorrect information to potential participants about the trial, describing it as smoking cessation support rather than research, which evidently impacted the expectations of some of the participants. This suggests more training may be required across all staff groups at sites.

#### 2. Evidence of intervention acceptability

Participants found the intervention appointment, which included communication with the clinician and the offer of an e-cigarette starter kit, acceptable. They felt that these components had a positive impact on their willingness and motivation to change their smoking behaviour. Researchers and clinicians also echoed that they felt that participants were generally satisfied with the intervention. Participants reported that they found the information leaflet useful but did not tend to look at it again after the initial appointment. Researchers also felt that the leaflet was comprehensive and explanatory enough for participants in this population.

#### 3. Improvements to the intervention

Clinicians felt that they did not have adequate time to deliver the intervention. Therefore, at one research site, research staff stepped in to do this and at another, researchers mentioned receiving a lot of questions about the e-cigarette afterwards. Participants with no e-cigarette experience felt that they required more assistance with getting to grips with the e-cigarette. This was also reported by two of the researchers. The findings suggest that on-going support/contact is therefore important, and participants also expressed that they would like ongoing support between intervention and follow-up. A sense of rejection and worry about not receiving an e-cigarette starter kit was mentioned a number of times. Researchers, clinicians, and participants reported that those allocated to the control group, who therefore did not receive an e-cigarette, felt disappointed. Additionally, most participants expressed worry that they would not receive an e-cigarette before randomization took place..

#### 4. Issues with the e-cigarette

Two participants believed that e-cigarettes are too expensive to continue after the trial finishes. The interviewed intervention participants mentioned some difficulties with the e-cigarette after leaving the appointment. One researcher reported that people did not seem to like tobacco-flavoured e-liquid.

#### Quantitative assessment of intervention acceptability

Table 2 shows that participants in the intervention provided high ratings for satisfaction of using the e-cigarettes, using e-cigarettes around friends and family, and highly rated the e-cigarette in terms of size, shape, feel and branding (all ratings >4 on 5-point scale). However, flavours were rated lower. One third of participants found the e-cigarette burdensome and nearly half reported a problem in using it. Both written and verbal information provided was generally deemed helpful and most participants said the intervention was the right length and two thirds of participants reported having positive experience taking part in the trial.

#### Mental health symptoms

There were no significant differences in changes in GAD-7 between baseline and follow-up between groups (Supplementary Table 3). For the changes in PHQ-9, the 80% CIs suggest a difference between groups for the change of item 9 between baseline and follow-up. There was a reduction in thoughts ‘that you would be better off dead or hurting yourself in some way’ in the intervention group but not in the control group. No other significant differences in PHQ-9 items were found (Supplementary Table 3).

#### Mood and physical symptoms, and adverse events

Constipation, irritable feeling and restlessness were higher in the intervention than control group (Supplementary Table 4). Overall adverse events in the past month at follow-up was low for both control and intervention group participants. However, the 80% CIs suggest a higher number of participants reporting nausea, irritation, depression, restlessness, increased appetite, dry mouth and throat, and wheezing in the intervention compared with the control group (Supplementary Table 4).

### Potential efficacy

#### CO validated sustained abstinence rates at 1-month follow-up

##### Planned analysis

No participants in the control group and 9.5% (n=2) in the experimental group reported CO validated sustained abstinence rates at 5 weeks with missing equalling smoking using intention to treat. Based on a confidence interval approach, this percentage difference (with upper 80% CI 15.0%) would suggest possible efficacy and evidence to proceed to the full trial.

In the complete case analysis, no participants in the control condition and 13.3% (n=2) in the experimental condition reported CO validated sustained abstinence rates at 5 weeks. Based on a confidence interval approach, this percentage difference (with upper 80% CI 21.1%) would suggest possible efficacy and evidence to proceed to the full trial.

##### Unplanned analysis

For the intention to treat analysis, the 90% confidence interval (-3.1% to 25.1%), 85% confidence interval (-0.9% to 22.7%) and 80% confidence interval (-0.7% to 20.9%) crossed both 0 and the clinically significant difference, this gives inconclusive evidence. The 75% confidence interval (1.9% to 19.4%) excludes 0 and crosses the clinically significant difference, at this level there is evidence of a treatment difference which is potentially clinically important. Only a 20% confidence interval or lower is wholly above or equal to a clinically significant difference, suggesting at this level that there is the clinically meaningful difference in smoking cessation between the groups.

For complete case analysis, the 90% confidence interval (-7.1% to 33.3%), 85% confidence interval (-3.5% to 28.2%), 80% confidence interval (-0.9%% to 28.2%) crossed both 0 and the clinically significant difference, this gives inconclusive evidence. The 75% confidence interval (0 excludes 0 and crosses the clinically significant difference, at this level there is evidence of a treatment difference which is potentially clinically important. Only a 45% confidence interval or lower is wholly above or equal to the clinically significant difference, suggesting at this level that there is a clinically meaningful difference in smoking cessation between the groups.

##### Point prevalence (24h) abstinence

In the intention to treat analysis with missing equal to smoking, 4.6% (n=1) of the participants in the control condition reported point prevalence abstinence, while 28.6% (n=6) in the experimental condition. This gives a percentage difference of 24.0% (with upper 80% CI of 33.3%). The 95% confidence interval (1.3% to 45.7%) excluded 0 indicating a difference between the two groups.

In the complete case analysis, 8.3% (n=1) of the participants in the control condition reported point prevalence abstinence, while 40.0% (n=6) in the experimental condition. This gives a percentage difference of 31.7% (with upper 80% CI of 43.5%). The 90% confidence interval (3.6% to 53.4%) excluded 0 indicating a difference between the two groups. The 95% confidence interval (-2.1% to 58.9%) did not excluded 0 indicating inconclusive evidence for a difference.

##### Smoking reduction

In the intention to treat analysis with missing equal to no change, 13.6% (n=3) of the participants in the control condition reported at least a 50% reduction in cigarette consumption, while 61.9% (n=13) in the experimental condition. This gives a percentage difference of 48.3% (with upper 80% CI of 58.1%). The 95% confidence interval (19.5% to 67.7%) excluded 0 indicating a difference between the two groups.

In the complete case analysis, 25.9% (n=3) of the participants in the control condition reported at least a 50% reduction in cigarette consumption, while 86.7% (n=13) in the experimental condition. This gives a percentage difference of 61.2% (with upper 80% CI of 72.2%). The 95% confidence interval (24.3% to 80.4%) excluded 0 indicating a difference between the two groups.

##### Set-up quit date and NRT use

At the 1-month follow-up, 77.8% (n=21) of the participants who were followed up had set a quit date; 93.3% (n=14) in the experimental condition and 58.3% (n=7) in the control condition. Among those who were followed up, 7.4% (n=2) reported using NRT in the past month, with one participant from each condition.

##### Health economics

Based on the instruments used in the feasibility study, the estimated cost per participant was £160 for the intervention group and £24 for the control group. For the intervention group, costs included £129 per participant for training, which covered the development of training materials, time spent by trainers and trainees, travel, and consumables. The cost of products for the trial was £11.48 for the device and £0.98 per bottle of e-liquid. Additionally, participants received a five-minute consultation with a clinician on using the e-cigarettes and a bespoke information leaflet, costing £8 to produce. The total mean intervention cost, excluding training expenses, was £31 per participant in the intervention group. For the control group, the mean cost of pharmacotherapies for smoking cessation was estimated at £4 per participant. The mean cost per participant for community smoking cessation aids, such as consultations with a GP or attending an NHS Stop Smoking Services session, was £20. Smokers in the control group incurred a mean cost of £24 per participant during the 1- month follow-up period. Supplementary Table 5 provides a detailed breakdown of intervention costs for both groups.

The health economics analysis confirms that it is possible to collect data from this population in preparation for a full RCT, including using a shortened version of the instruments employed.

## Discussion

This feasibility RCT aimed to assess the feasibility and acceptability of providing an e-cigarette starter kit (with additional support on how to use the e-cigarettes) to smokers with mental health condition as an adjunct to their usual care in UK primary and secondary care settings. While the target sample size was not achieved, raising questions over the feasibility of our approach, our findings provide preliminary evidence that the trial and intervention were broadly acceptable to participants and health professionals delivering the treatment, well tolerated, achieving good consenting and adherence rates, with limited contamination. In addition, our exploration of preliminary effect size indicated potential efficacy, as continuous and point prevalence abstinence and reductions in cigarette consumption were more pronounced in the intervention than control group. However, event rates were low, which reduced the precision of estimates, and different confidence intervals were calculated to determine the level at which a treatment effect might be present The cost of the intervention was in line with similar smoking cessation treatments in this population (49).

As expected, our sample scored moderately to highly on measures of anxiety and depression, though GAD-7 and PHQ-9 scores remained stable or declined slightly from baseline to follow-up, suggesting that mental health condition does not necessarily worsen during a quit attempt and may potentially improve. However, given the small sample size and the short follow-up period of one month, where withdrawal symptoms may still be present, these findings should be interpreted with caution. Such findings, though, align with previous research which has demonstrated that stopping smoking is associated with an improvement in mental health condition symptoms (4). Health professionals working with people with mental health conditions are often concerned about worsening mental health outcomes, and this has been a key barrier to both starting discussions around smoking behaviour change, and also implementing smoking cessation programmes for this population (50). While health professionals should find this growing body of evidence reassuring, indicating that smoking cessation does not counter progress with other mental health symptoms in adults with mental health conditions, further research with larger sample sizes and longer follow-up periods is needed to confirm these findings.

This feasibility study also highlighted some problems in terms of the practicality of conducting a trial within community mental health teams and associated research procedures, failing to achieve the desired recruitment rate, with attrition higher than other smoking cessation studies in this or similar populations (16, 19, 33). Quantitative analysis and the qualitative interviews provided further insights into potential barriers to undertaking a full RCT to evaluate the intervention in this setting.

In terms of study processes and design, recruitment was the biggest challenge. The number of eligible participants varied considerably across sites, with some having small caseloads per clinician and low numbers of new referrals, meaning recruitment sources were exhausted quickly. There were numerous sites where clinicians had no availability to book in trial participants for several weeks, while some patients at the involved sites were only seen once per year by their clinicians and did not appear to have the commonly expected engagement with services, or rapport with the clinicians. The latter also could have impacted the attrition rate. To address these issues, several mitigation strategies to improve recruitment have been proposed (Supplementary Table 6), based on review of previous research (33, 51, 52). In addition, more realistic recruitment targets may need to be adopted for future trials in this setting.

Lastly, attrition was higher than expected. Participants reported being disappointed and feeling negatively affected in terms of their mental health when learning they had been randomised to the control group. We are mindful that many patients in our population may have tried to quit smoking unsuccessfully for a long time and may therefore be hopeful to receive the active intervention, especially the e-cigarette device itself. Consequently, it would be equitable to offer control group participants the e-cigarette kit for free together with a video link and a leaflet at their last follow-up appointment to address disappointment and attrition in this group, which was higher than the intervention group as this would also serve as an additional incentive.

In terms of intervention content and delivery, the qualitative data indicated that some patients found the technicalities involved in using the e-cigarette challenging and would have appreciated further support. Additionally, the intervention group had higher mood and physical symptom scores compared to the control group. These differences may be due to vaping and the process of smoking cessation or switching from cigarettes to e-cigarettes. This aligns with previous research indicating that smokers often experience adverse side effects from vaping (53). In light of this, it would be preferrable for future work with this population to move from a tank-based model (which was the only evidence-based model at the time of the project proposal) to a pod-based model, as this has the advantage of being substantially easier to use. It is also tamper-proof and thus offers fewer opportunities for potential misuse. Additionally, unpublished consumer research by the University of East Anglia, conducted as part of a trial assessing the effectiveness of e-cigarette provision in emergency care settings, showed that a pod device received high ratings for ease of use and satisfaction (52, 54). Further, the qualitative findings indicated that five minutes of time factored in for the delivery of the intervention was insufficient. Future studies should therefore consider allowing more time for the delivery of brief interventions, given these particular challenges and to offer additional support (e.g., a ‘telephone helpline’ staffed by our researchers). Relatedly, if pod instead of tank devices are used, this likely will free up time, as these are easier to use, thus can be demonstrated more quickly, and simpler, thus requiring less additional support.

## Strengths and Limitations

To our knowledge, this is the first study in the UK to explore the feasibility and acceptability of supplying an e-cigarette starter kit as a long-term harm reduction and smoking cessation tool to people with mental health condition treated in the community as adjunct to usual care. This study used robust quantitative and qualitative methodology to evaluate the feasibility of delivering the intervention, which was well received with minimal negative effects. However, there were several limitations. First, we did not meet our original target recruitment rate, which affected our ability to draw firm conclusion about potential efficacy. Second, and relatedly, recruitment and attrition rate posed challenges and differed substantially across sites. This is an important finding, which will assist us in carefully selecting suitable sites in a future main trial. Third, event rates were low, reducing precision of estimates. Fourth, follow-up rates differed between treatment groups. However, findings from complete case analysis were consistent with the primary analysis. Fifth, the assessment of adherence to treatment was based on a crude measure of the proportion of participants who were using an e-cigarette at 1-month follow-up. We will consider a more rigorous assessment for the full trial. Sixth, even though the mental health diagnoses of participants were taken from their health records, we were not able to record the exact mental health diagnosis of some participants. Our difficulties to obtain standard baseline data relating to participants’ diagnoses despite exhaustive attempts involving all relevant sites indicate that researchers may want to be mindful of practical difficulties possibly caused by staffing and capacity issues in UK NHS mental health contexts post Covid.

## Conclusions

Overall, findings from this feasibility study demonstrate that offering an intervention comprising of an e-cigarette starter kit, brief demonstration, verbal and written advice to smokers with mental health condition treated in the community is broadly acceptable and may be beneficial for participants in terms of harm reduction outcome such as switching from cigarette smoking to e-cigarette use and substantial reduction in cigarette consumption, with minimal negative effects. Findings also identified a number of barriers to undertaking a trial in this setting. These insights can be used to inform the design of future harm reduction trials in similar contexts.

## Data Availability

The datasets used and/or analysed during the current study are available from the corresponding author on reasonable request.

## List of abbreviation

NRT: Nicotine Replacement Therapy
RCT: Randomised Controlled Trial
OR: Odds Ratio
CI: Confidence Intervals
SD: Standard Deviation

## Ethics approval and consent to participate

Ethical approval for the study was granted by National Health System UK (NHS) Health Research Authority (REC ref:21/NE/0202). Informed consent was obtained from all participants.

## Consent for publication

Not applicable.

## CrediT authorship contribution statement

DK: Writing – original draft, Writing – review & editing; EB: Formal analysis; Writing – review & editing; AM: Data curation; Formal analysis; Writing – review & editing; JP: Formal analysis; Writing – review & editing; QW: Formal analysis; Writing – review & editing; ER: Conceptualization, Writing – review & editing, Funding acquisition; LS: Conceptualization, Writing – review & editing, Funding acquisition.

## Supplementary material

**Supplementary Table 1.**
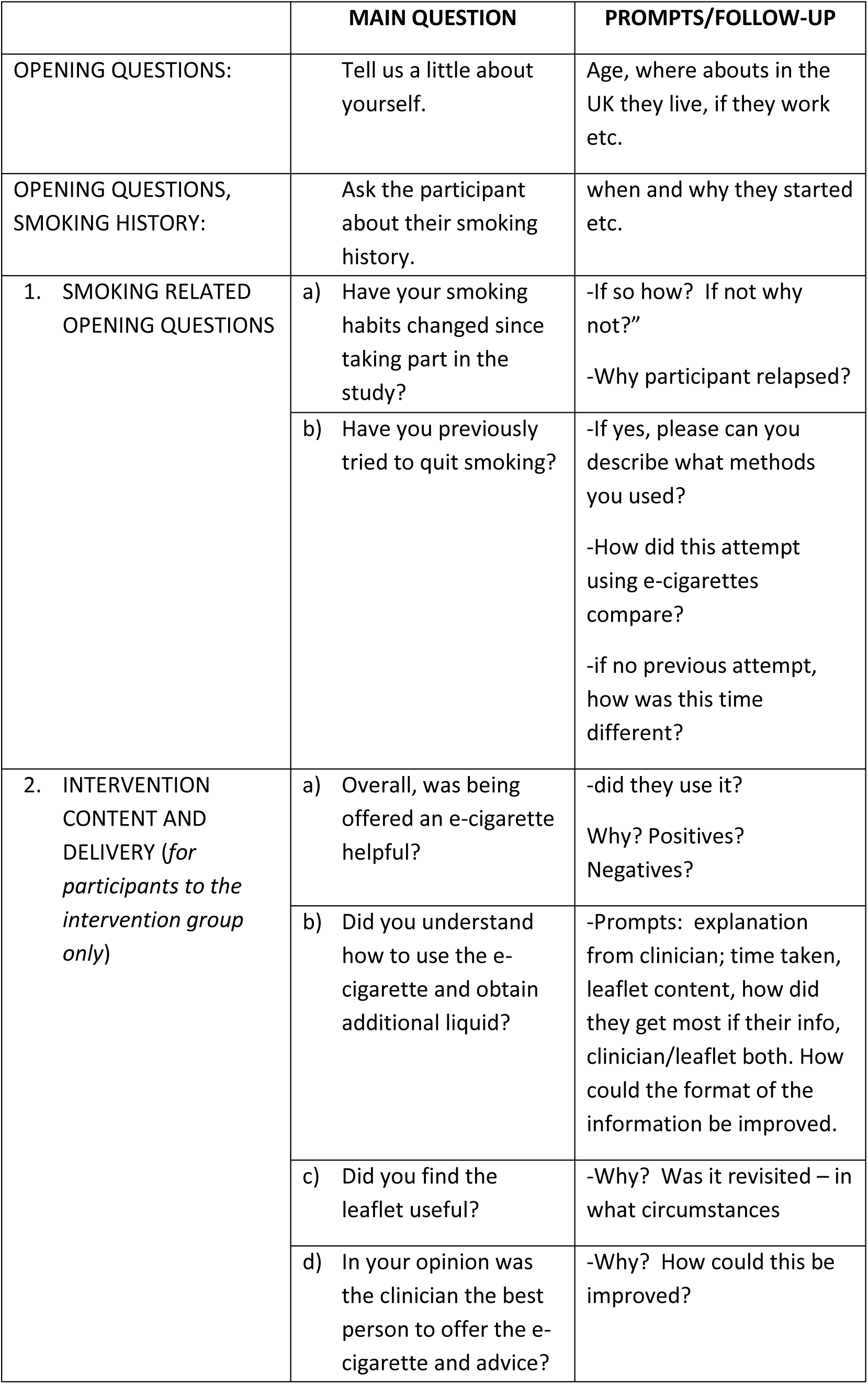
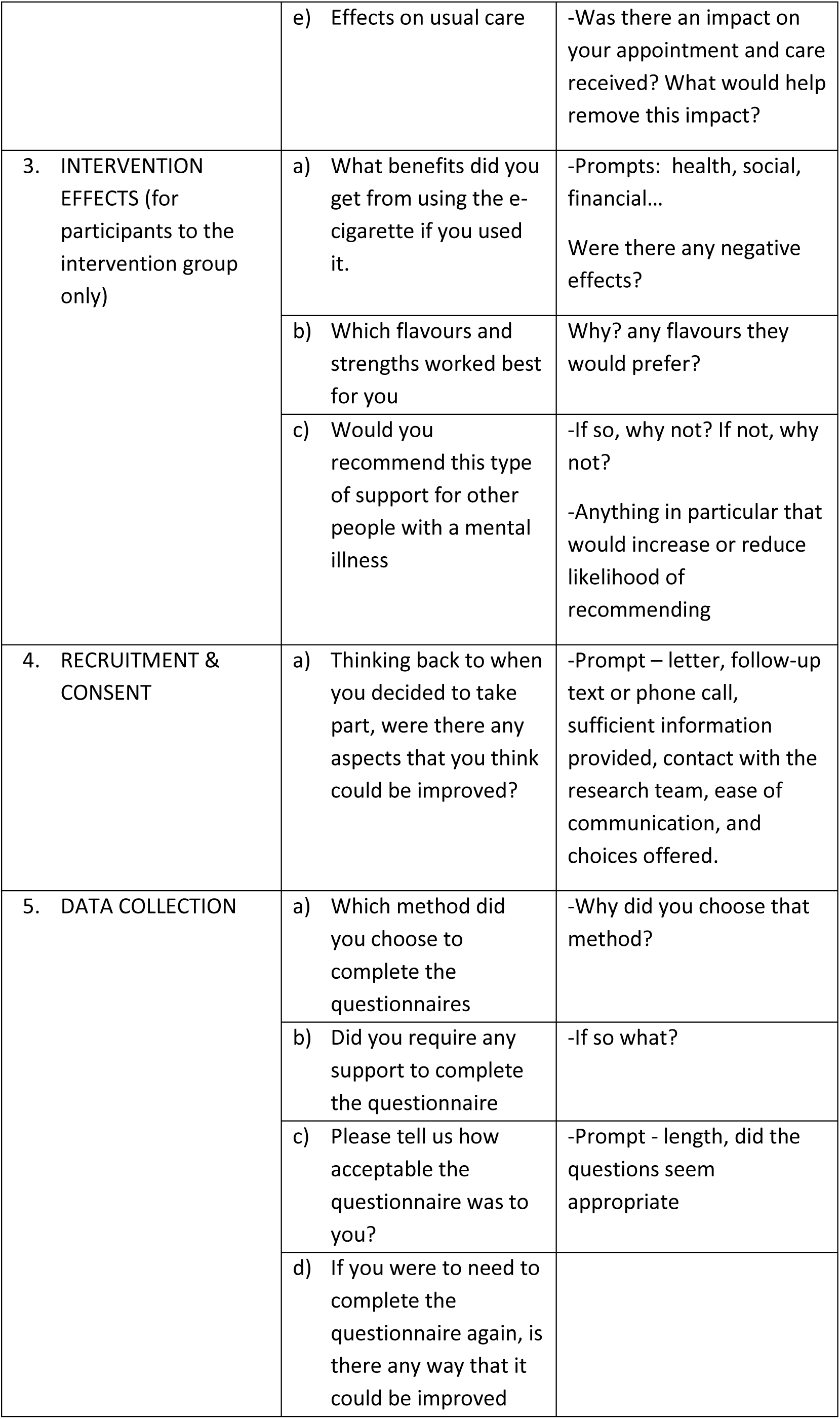
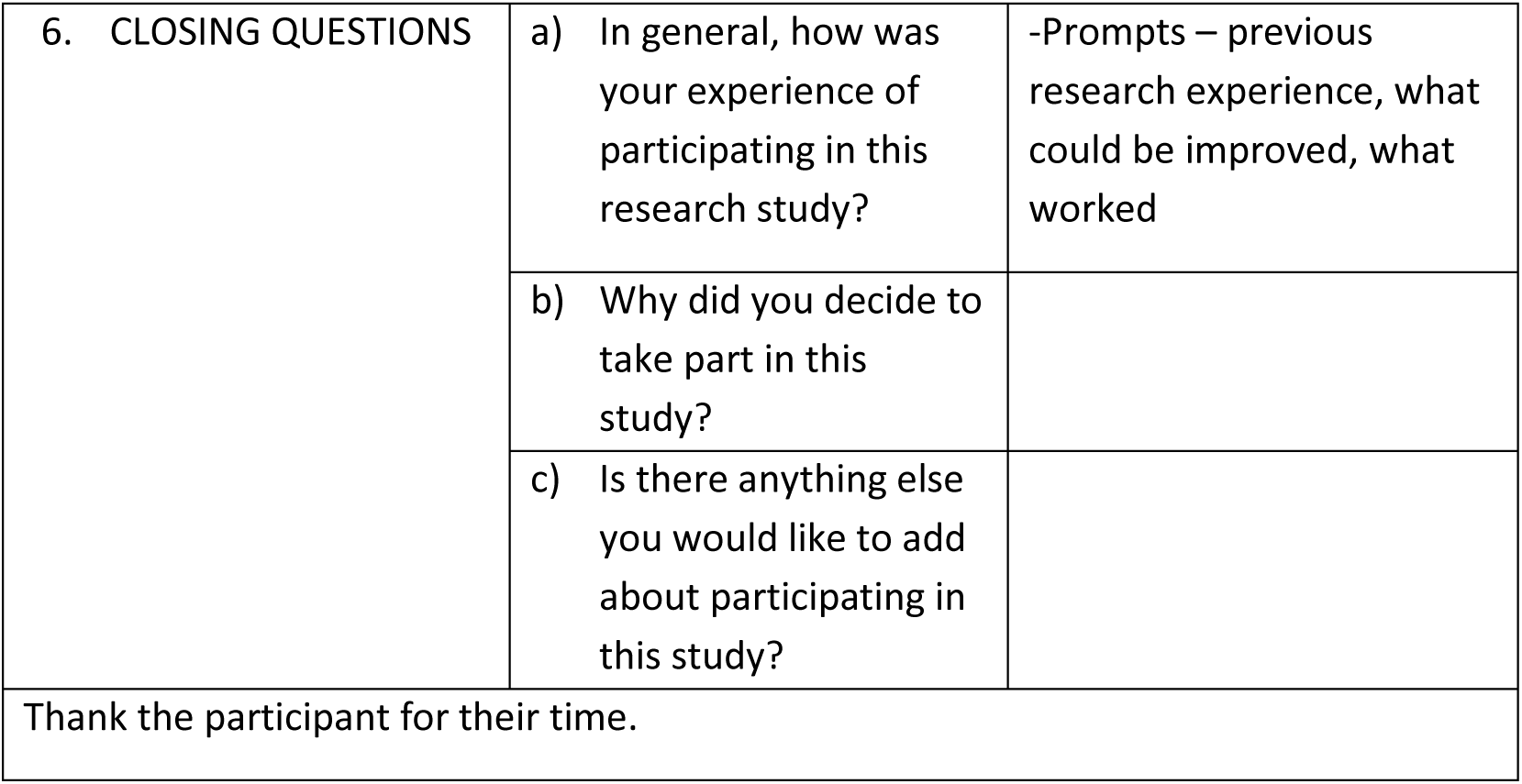
Interview topic guide.

|  | MAIN QUESTION | PROMPTS/FOLLOW-UP |
| --- | --- | --- |
| OPENING QUESTIONS: | Tell us a little about yourself. | Age, whereabouts in the UK they live, if they work etc. |
| OPENING QUESTIONS, SMOKING HISTORY: | Ask the participant about their smoking history. | when and why they started etc. |
| 1. SMOKING RELATED OPENING QUESTIONS | a) Have your smoking habits changed since taking part in the study? | -If so how? If not why not?"<br>-Why participant relapsed? |
|  | b) Have you previously tried to quit smoking? | -If yes, please can you describe what methods you used?<br>-How did this attempt using e-cigarettes compare?<br>-if no previous attempt, how was this time different? |
| 2. INTERVENTION CONTENT AND DELIVERY (*for participants to the intervention group only*) | a) Overall, was being offered an e-cigarette helpful? | -did they use it?<br>Why? Positives?<br>Negatives? |
|  | b) Did you understand how to use the e-cigarette and obtain additional liquid? | -Prompts: explanation from clinician; time taken, leaflet content, how did they get most of their info, clinician/leaflet both. How could the format of the information be improved. |
|  | c) Did you find the leaflet useful? | -Why? Was it revisited – in what circumstances |
|  | d) In your opinion was the clinician the best person to offer the e-cigarette and advice? | -Why? How could this be improved? |
|  | e) Effects on usual care | -Was there an impact on your appointment and care received? What would help remove this impact? |
| 3. INTERVENTION EFFECTS (for participants to the intervention group only) | a) What benefits did you get from using the e-cigarette if you used it. | -Prompts: health, social, financial...<br>Were there any negative effects? |
|  | b) Which flavours and strengths worked best for you | Why? any flavours they would prefer? |
|  | c) Would you recommend this type of support for other people with a mental illness | -If so, why not? If not, why not?<br>-Anything in particular that would increase or reduce likelihood of recommending |
| 4. RECRUITMENT & CONSENT | a) Thinking back to when you decided to take part, were there any aspects that you think could be improved? | -Prompt – letter, follow-up text or phone call, sufficient information provided, contact with the research team, ease of communication, and choices offered. |
| 5. DATA COLLECTION | a) Which method did you choose to complete the questionnaires | -Why did you choose that method? |
|  | b) Did you require any support to complete the questionnaire | -If so what? |
|  | c) Please tell us how acceptable the questionnaire was to you? | -Prompt - length, did the questions seem appropriate |
|  | d) If you were to need to complete the questionnaire again, is there any way that it could be improved |  |
| 6. CLOSING QUESTIONS | a) In general, how was your experience of participating in this research study? | -Prompts – previous research experience, what could be improved, what worked |
|  | b) Why did you decide to take part in this study? |  |
|  | c) Is there anything else you would like to add about participating in this study? |  |
| Thank the participant for their time. |  |  |

**Supplementary Table 2:**
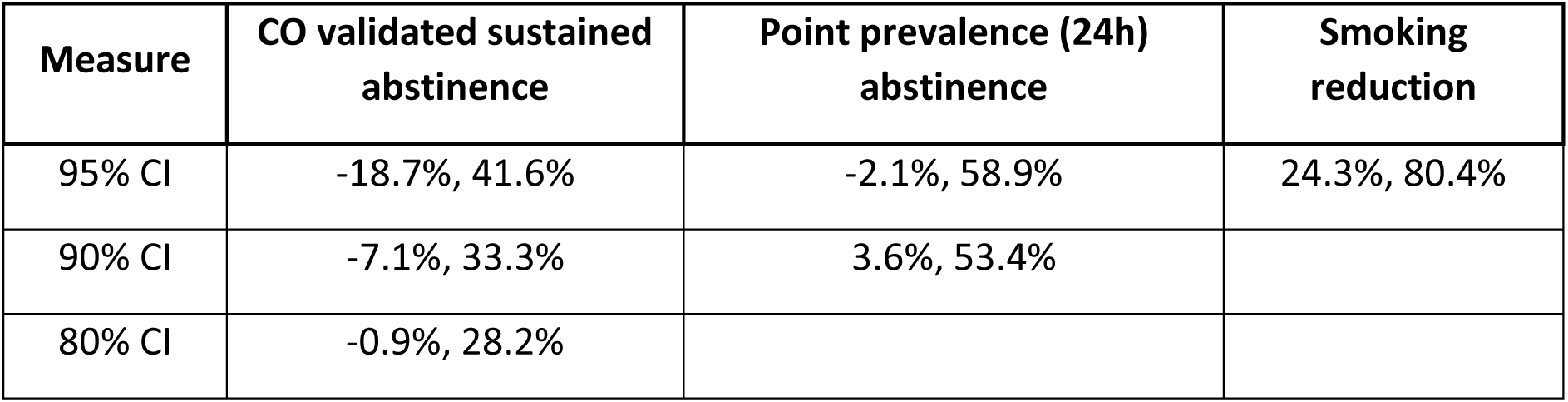
Group differences with scaled confidence intervals (complete case analysis)

| Measure | CO validated sustained abstinence | Point prevalence (24h) abstinence | Smoking reduction |
| --- | --- | --- | --- |
| 95% CI | -18.7%, 41.6% | -2.1%, 58.9% | 24.3%, 80.4% |
| 90% CI | -7.1%, 33.3% | 3.6%, 53.4% |  |
| 80% CI | -0.9%, 28.2% |  |  |

**Supplementary Table 3:**
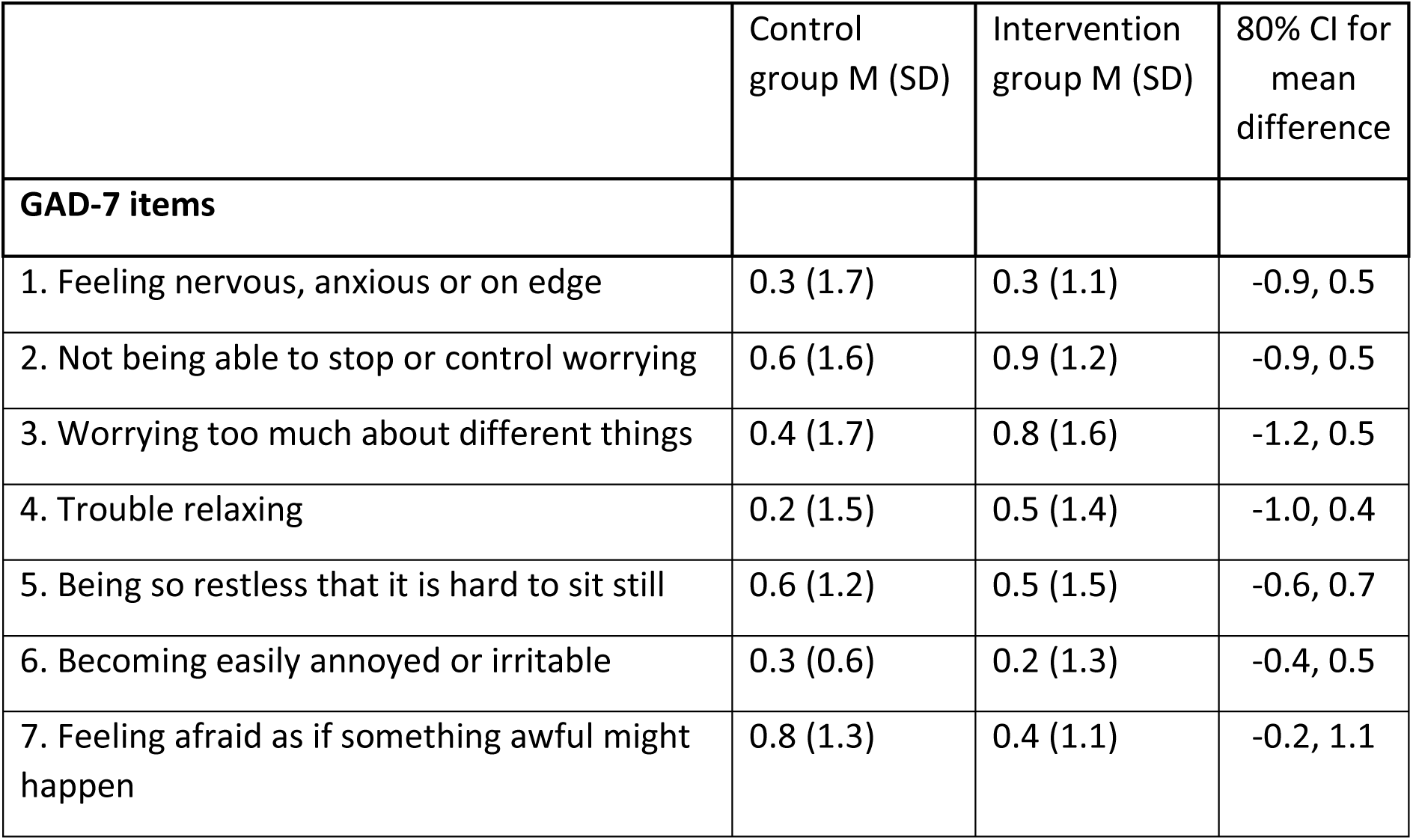
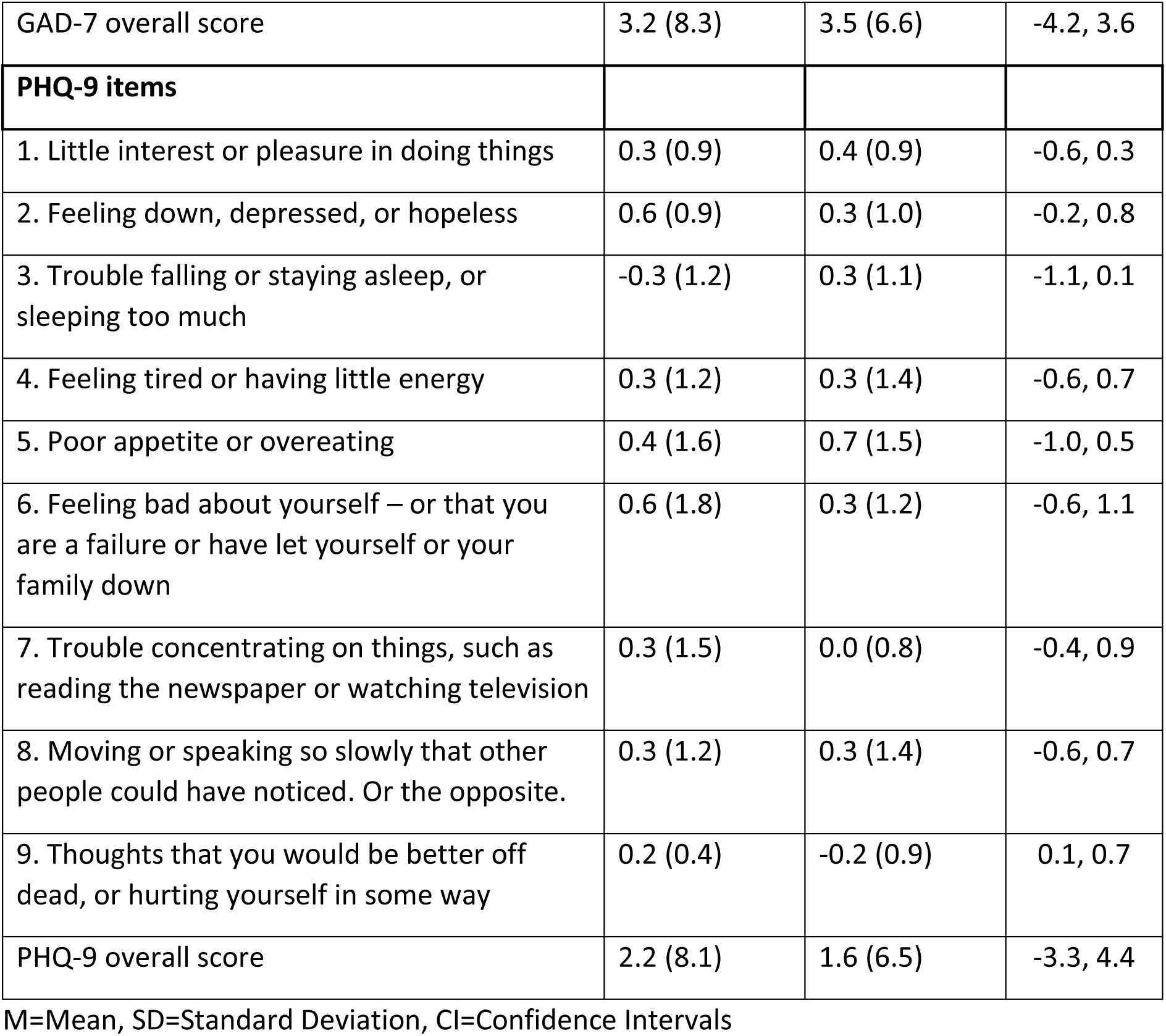
Changes in GAD-7 and PHQ-9 items between baseline and follow-up.

|  | Control group M (SD) | Intervention group M (SD) | 80% CI for mean difference |
| --- | --- | --- | --- |
| **GAD-7 items** |  |  |  |
| 1. Feeling nervous, anxious or on edge | 0.3 (1.7) | 0.3 (1.1) | -0.9, 0.5 |
| 2. Not being able to stop or control worrying | 0.6 (1.6) | 0.9 (1.2) | -0.9, 0.5 |
| 3. Worrying too much about different things | 0.4 (1.7) | 0.8 (1.6) | -1.2, 0.5 |
| 4. Trouble relaxing | 0.2 (1.5) | 0.5 (1.4) | -1.0, 0.4 |
| 5. Being so restless that it is hard to sit still | 0.6 (1.2) | 0.5 (1.5) | -0.6, 0.7 |
| 6. Becoming easily annoyed or irritable | 0.3 (0.6) | 0.2 (1.3) | -0.4, 0.5 |
| 7. Feeling afraid as if something awful might happen | 0.8 (1.3) | 0.4 (1.1) | -0.2, 1.1 |
| GAD-7 overall score | 3.2 (8.3) | 3.5 (6.6) | -4.2, 3.6 |
| **PHQ-9 items** |  |  |  |
| 1. Little interest or pleasure in doing things | 0.3 (0.9) | 0.4 (0.9) | -0.6, 0.3 |
| 2. Feeling down, depressed, or hopeless | 0.6 (0.9) | 0.3 (1.0) | -0.2, 0.8 |
| 3. Trouble falling or staying asleep, or sleeping too much | -0.3 (1.2) | 0.3 (1.1) | -1.1, 0.1 |
| 4. Feeling tired or having little energy | 0.3 (1.2) | 0.3 (1.4) | -0.6, 0.7 |
| 5. Poor appetite or overeating | 0.4 (1.6) | 0.7 (1.5) | -1.0, 0.5 |
| 6. Feeling bad about yourself – or that you are a failure or have let yourself or your family down | 0.6 (1.8) | 0.3 (1.2) | -0.6, 1.1 |
| 7. Trouble concentrating on things, such as reading the newspaper or watching television | 0.3 (1.5) | 0.0 (0.8) | -0.4, 0.9 |
| 8. Moving or speaking so slowly that other people could have noticed. Or the opposite. | 0.3 (1.2) | 0.3 (1.4) | -0.6, 0.7 |
| 9. Thoughts that you would be better off dead, or hurting yourself in some way | 0.2 (0.4) | -0.2 (0.9) | 0.1, 0.7 |
| PHQ-9 overall score | 2.2 (8.1) | 1.6 (6.5) | -3.3, 4.4 |

M=Mean, SD=Standard Deviation, CI=Confidence Intervals

**Supplementary Table 4:**
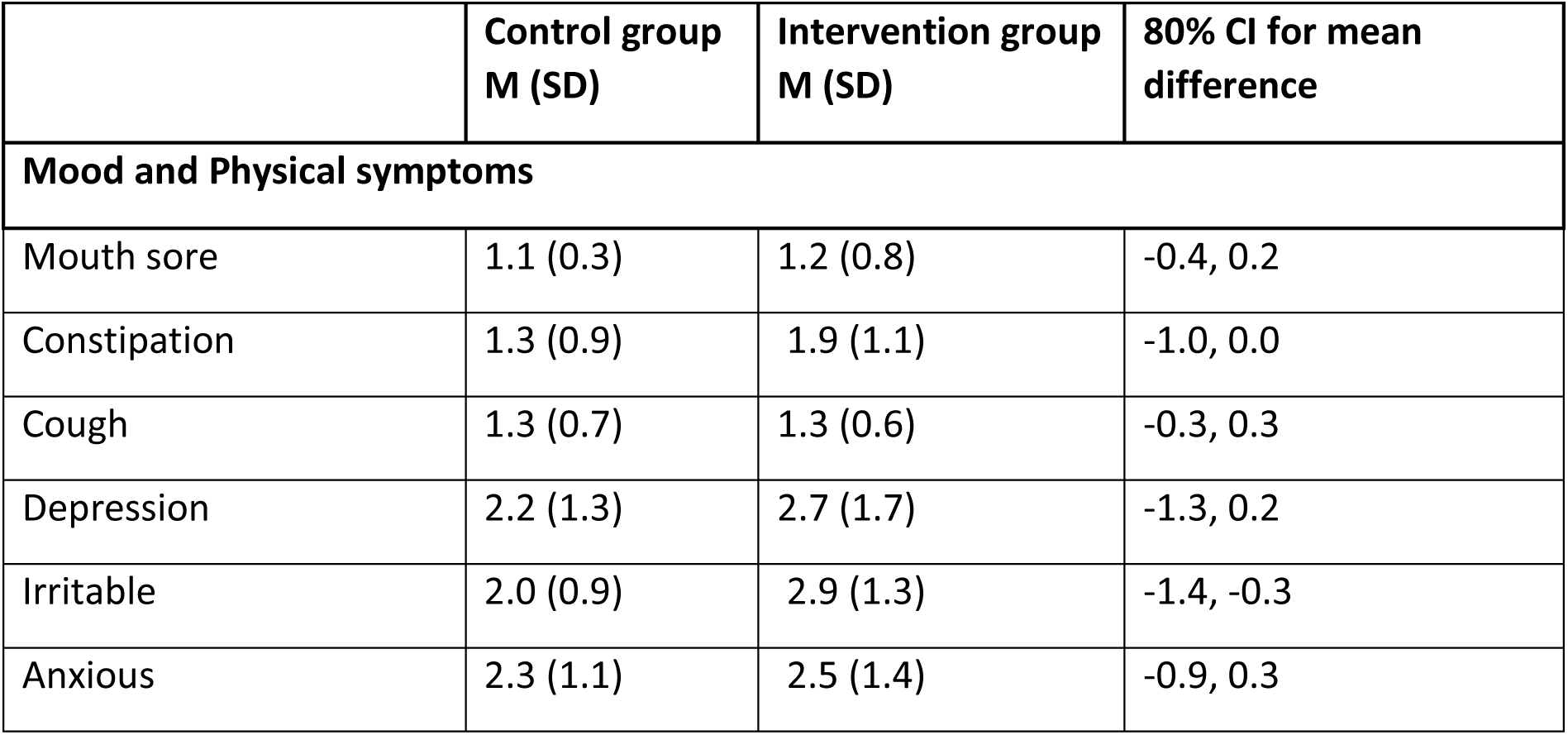
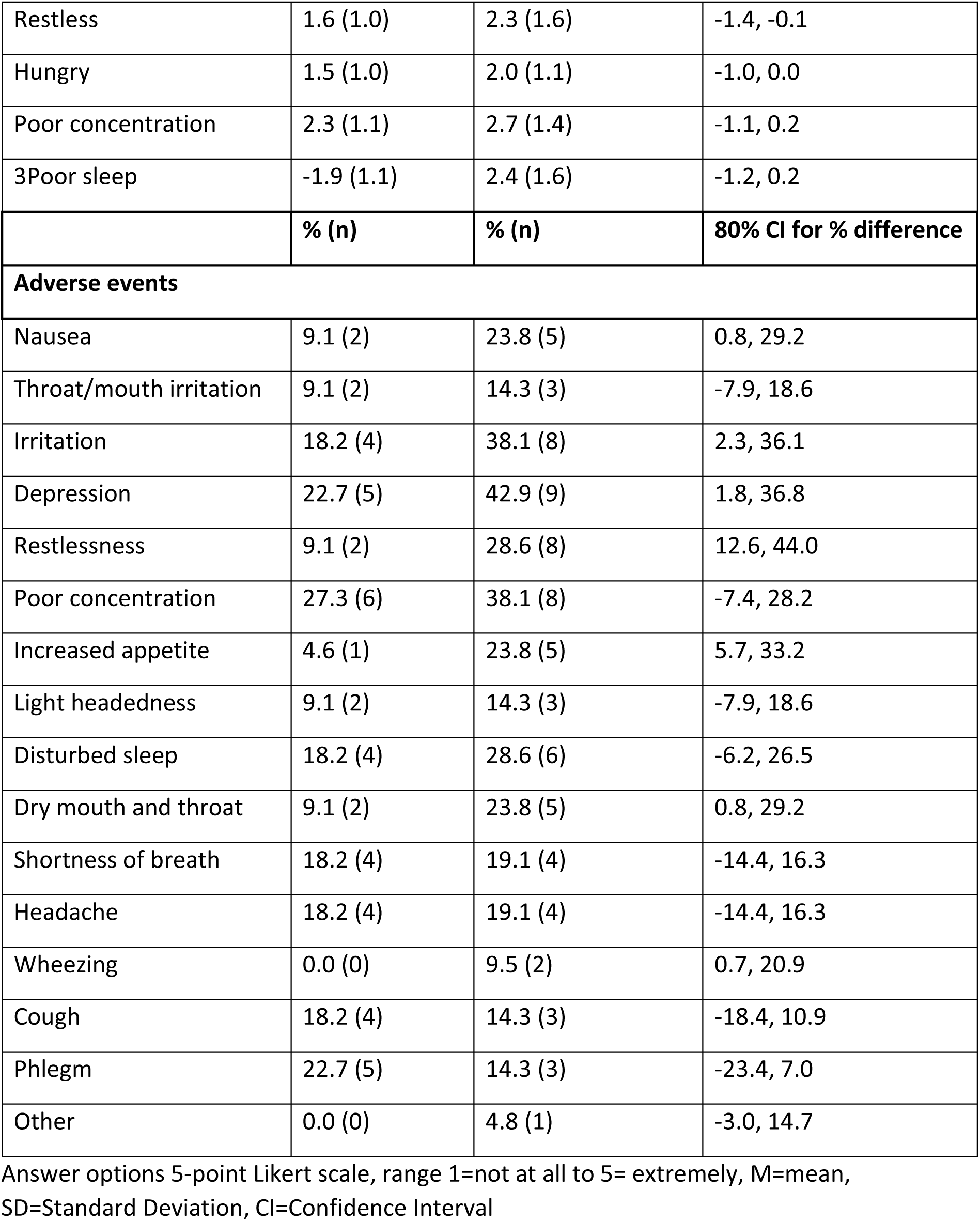
General mood and physical symptoms and adverse events as function of group assignment.

**Supplementary Table 5.** Breakdown of intervention cost.

| Cost component | Cost per participant |
| --- | --- |
| Intervention group |  |
| Training |  |
| Preparation of training materials | £17 |
| Staff cost (trainer) for face-to-face sessions | £29 |
| Intervention group |  |
| Training |  |
| Staff cost (trainees) for both face-to-face training and NCSTS online training | £82 |
| Consumables | £1 |
| Total training cost | £129 |
| Brief consultation | £3 |
| Bespoke information leaflet | £8 |
| e-cigarette |  |
| Device | £11 |
| e-liquid | £9 |
| Total intervention cost (without training) | £31 |
| Total intervention cost (with training) | £160 |
| Control group |  |
| Pharmacotherapies for smoking cessation | £4 |
| Primary care related to smoking cessation | £20 |
| Total smoking cessation cost | £24 |

**Supplementary Table 6.**
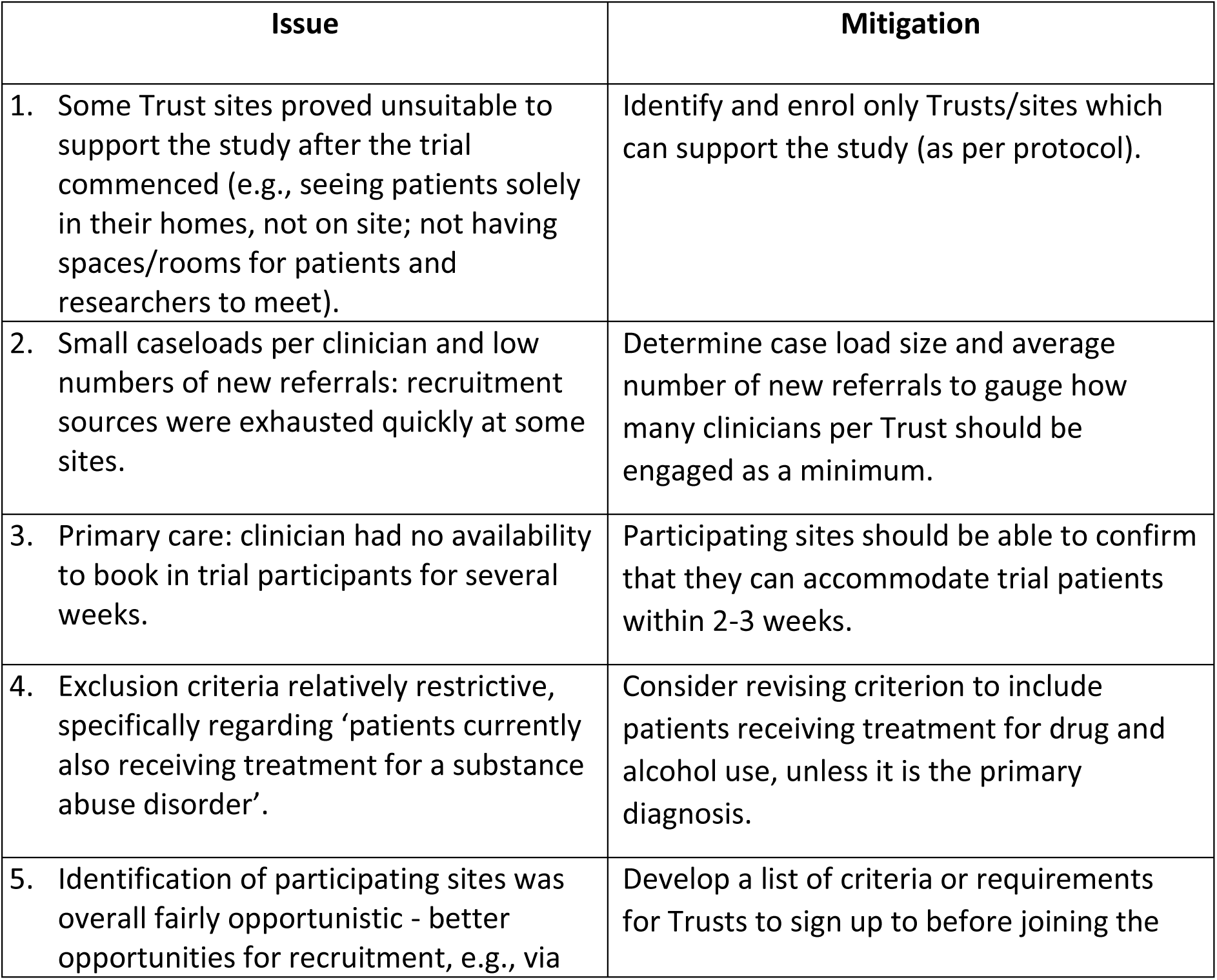
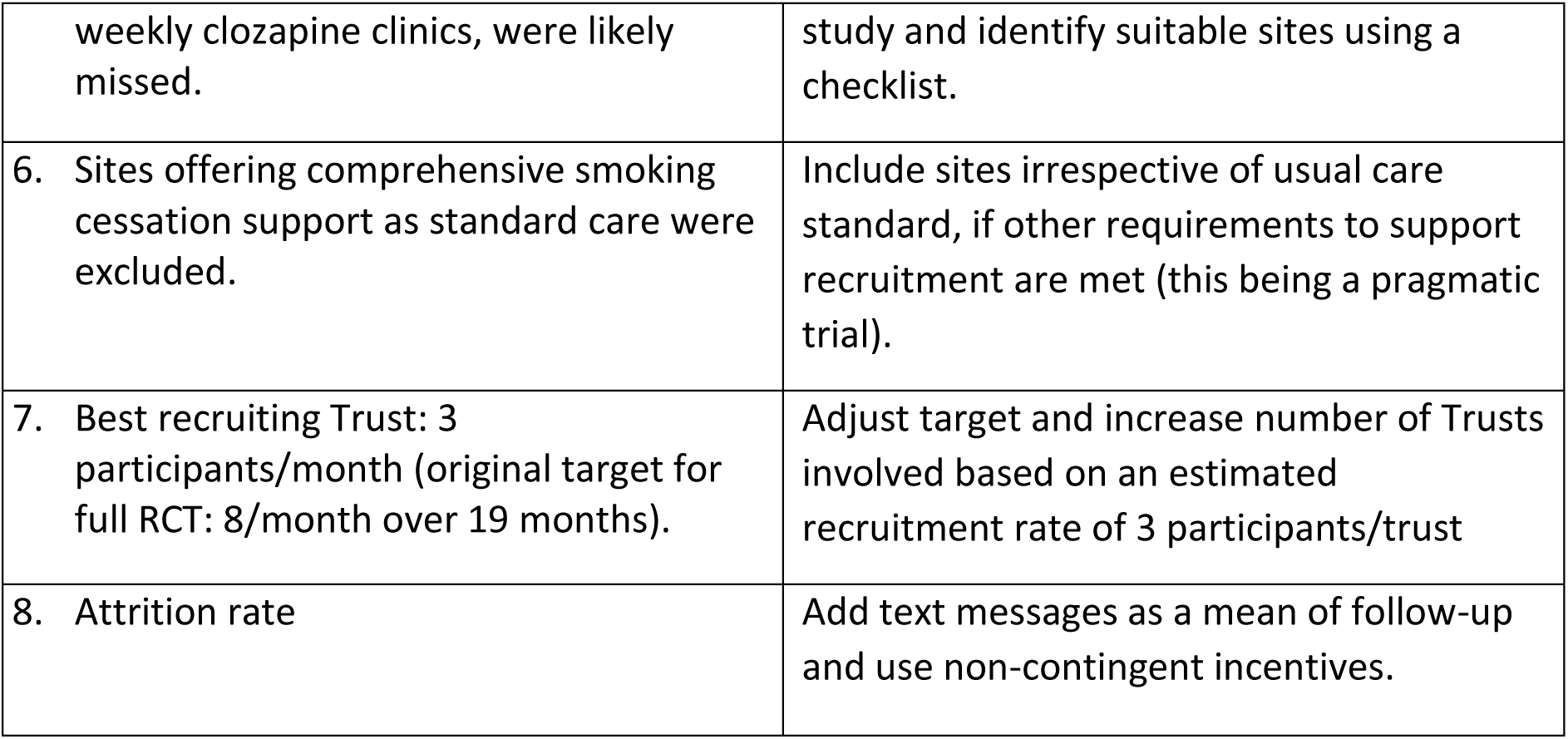
Recruitment issues and mitigation strategies.

CONSORT checklist of information to include when reporting a pilot or feasibility trial
| Section/Topic | Item No | Checklist item | Reported on page No |
| --- | --- | --- | --- |
| **Title and abstract** |  |  |  |
|  | 1a | Identification as a pilot or feasibility randomised trial in the title | 1 |
|  | 1b | Structured summary of pilot trial design, methods, results, and conclusions (for specific guidance see CONSORT abstract extension for pilot trials) | 2-3 |
| **Introduction** |  |  |  |
| Background and objectives | 2a | Scientific background and explanation of rationale for future definitive trial, and reasons for randomised pilot trial | 4-5 |
|  | 2b | Specific objectives or research questions for pilot trial | 5-6 |
| **Methods** |  |  |  |
| Trial design | 3a | Description of pilot trial design (such as parallel, factorial) including allocation ratio | 6 |
|  | 3b | Important changes to methods after pilot trial commencement (such as eligibility criteria), with reasons | n/a |
| Participants | 4a | Eligibility criteria for participants | 7 |
|  | 4b | Settings and locations where the data were collected | 7 |
|  | 4c | How participants were identified and consented | 6-7 |
| Interventions | 5 | The interventions for each group with sufficient details to allow replication, including how and when they were actually administered | 8-9 |
| Outcomes | 6a | Completely defined pre-specified assessments or measurements to address each pilot trial objective specified in 2b, including how and when they were assessed | 10-13 |
|  | 6b | Any changes to pilot trial assessments or measurements after the pilot trial commenced, with reasons | n/a |
|  | 6c | If applicable, prespecified criteria used to judge whether, or how, to proceed with future definitive trial | 10-13 |
| Sample size | 7a | Rationale for numbers in the pilot trial | 13 |
|  | 7b | When applicable, explanation of any interim analyses and stopping guidelines | n/a |
| Randomisation: |  |  |  |
| Sequence generation | 8a | Method used to generate the random allocation sequence | 7-8 |
|  | 8b | Type of randomisation(s); details of any restriction (such as blocking and block size) | 7-8 |
| Allocation concealment mechanism | 9 | Mechanism used to implement the random allocation sequence (such as sequentially numbered containers), describing any steps taken to conceal the sequence until interventions were assigned | 7-8 |
| Implementation | 10 | Who generated the random allocation sequence, who enrolled participants, and who assigned participants to interventions | 6-7 |
| Blinding | 11a | If done, who was blinded after assignment to interventions (for example, participants, care providers, those assessing outcomes) and how | 8 |
|  | 11b | If relevant, description of the similarity of interventions | 7-8 |
| Statistical methods | 12 | Methods used to address each pilot trial objective whether qualitative or quantitative | 12-13 |
| **Results** |  |  |  |
| Participant flow (a diagram is strongly recommended) | 13a | For each group, the numbers of participants who were approached and/or assessed for eligibility, randomly assigned, received intended treatment, and were assessed for each objective | 34. figure 1 |
|  | 13b | For each group, losses and exclusions after randomisation, together with reasons | 16 |
| Recruitment | 14a | Dates defining the periods of recruitment and follow-up | 6, 9 |
|  | 14b | Why the pilot trial ended or was stopped | n/a |
| Baseline data | 15 | A table showing baseline demographic and clinical characteristics for each group | 34, Table 1 |
| Numbers analysed | 16 | For each objective, number of participants (denominator) included in each analysis. If relevant, these numbers should be by randomised group | 15-22 |
| Outcomes and estimation | 17 | For each objective, results including expressions of uncertainty (such as 95% confidence interval) for any estimates. If relevant, these numbers should be by randomised group | 15-22 |
| Ancillary analyses | 18 | Results of any other analyses performed that could be used to inform the future definitive trial | 15-22 |
| Harms | 19 | All important harms or unintended effects in each group (for specific guidance see CONSORT for harms) | 19 |
|  | 19a | If relevant, other important unintended consequences | n/a |
| **Discussion** |  |  |  |
| Limitations | 20 | Pilot trial limitations, addressing sources of potential bias and remaining uncertainty about feasibility | 25 |
| Generalisability | 21 | Generalisability (applicability) of pilot trial methods and findings to future definitive trial and other studies | 23-25 |
| Interpretation | 22 | Interpretation consistent with pilot trial objectives and findings, balancing potential benefits and harms, and considering other relevant evidence | 22-25 |
|  | 22a | Implications for progression from pilot to future definitive trial, including any proposed amendments | 23-25 |
| **Other information** |  |  |  |
| Registration | 23 | Registration number for pilot trial and name of trial registry | 3 |
| Protocol | 24 | Where the pilot trial protocol can be accessed, if available | n/a |
| Funding | 25 | Sources of funding and other support (such as supply of drugs), role of funders | 26 |
|  | 26 | Ethical approval or approval by research review committee, confirmed with reference number | 6, 26 |

\*We strongly recommend reading this statement in conjunction with the CONSORT 2010 Explanation and Elaboration for important clarifications on all the items. If relevant, we also recommend reading CONSORT extensions for cluster randomised trials, non-inferiority and equivalence trials, non-pharmacological treatments, herbal interventions, and pragmatic trials. Additional extensions are forthcoming: for those and for up to date references relevant to this checklist, see www.consort-statement.org.

**Supplementary Table 7.**
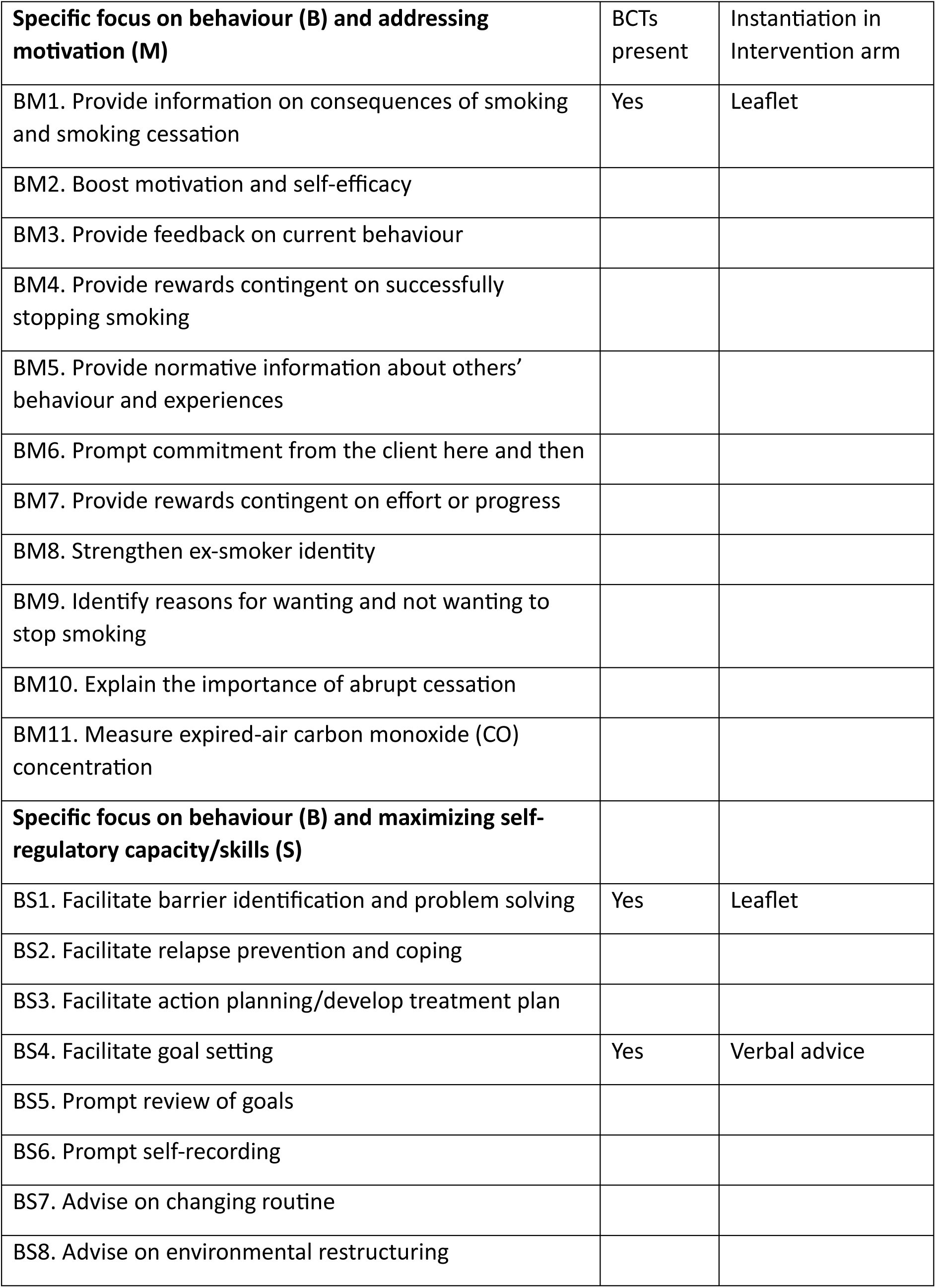
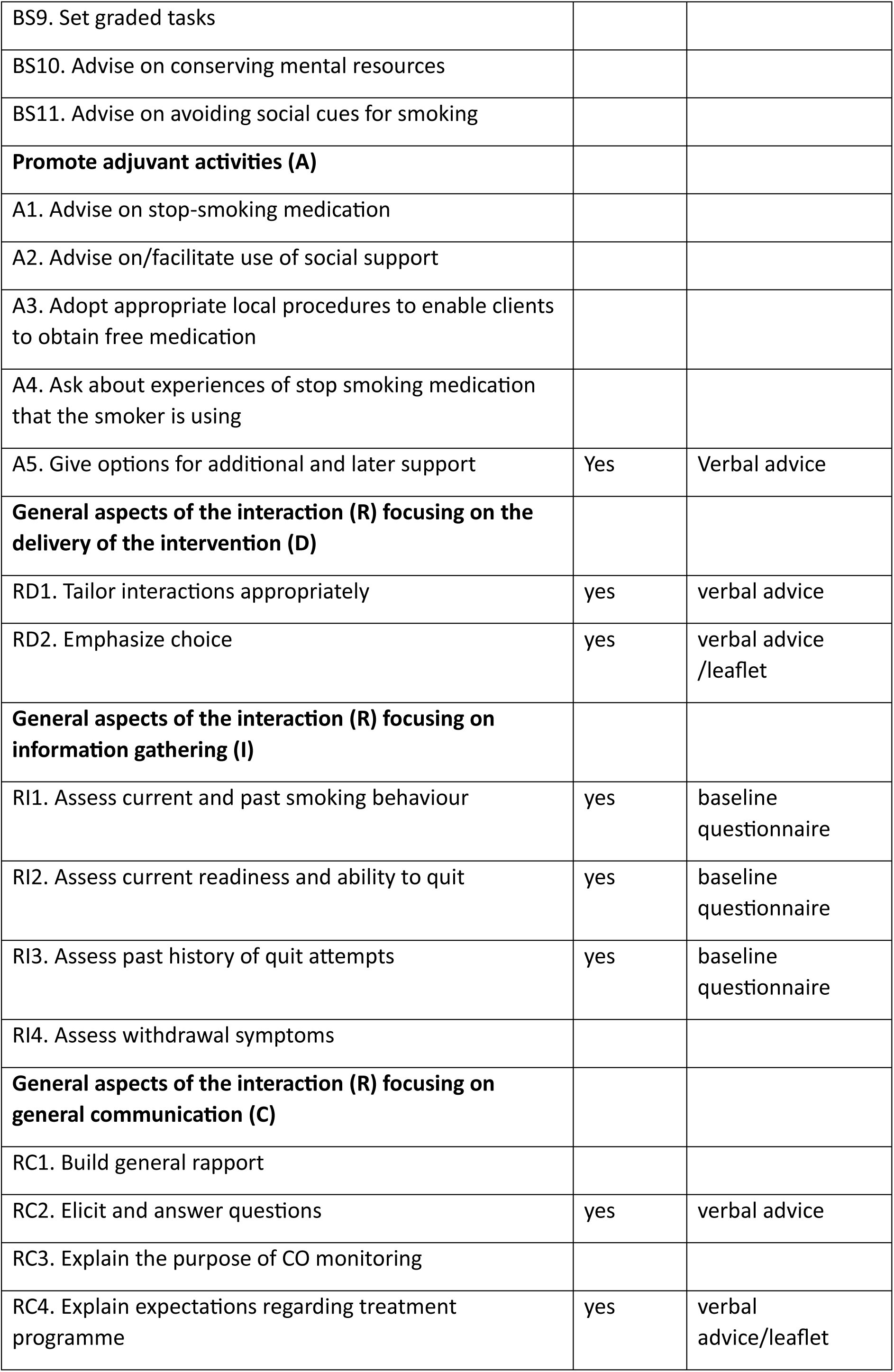
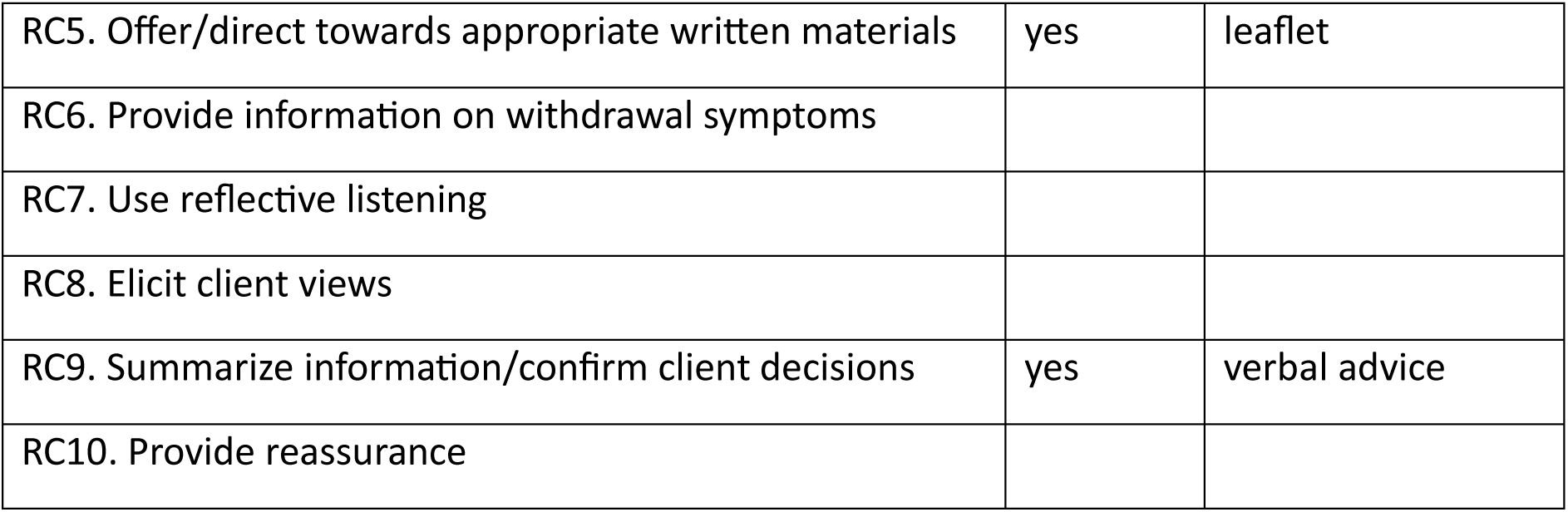
Behaviour change techniques (BCTs) included in the intervention, coded against a 44-item taxonomy of BCTs used in behavioural smoking cessation interventions.

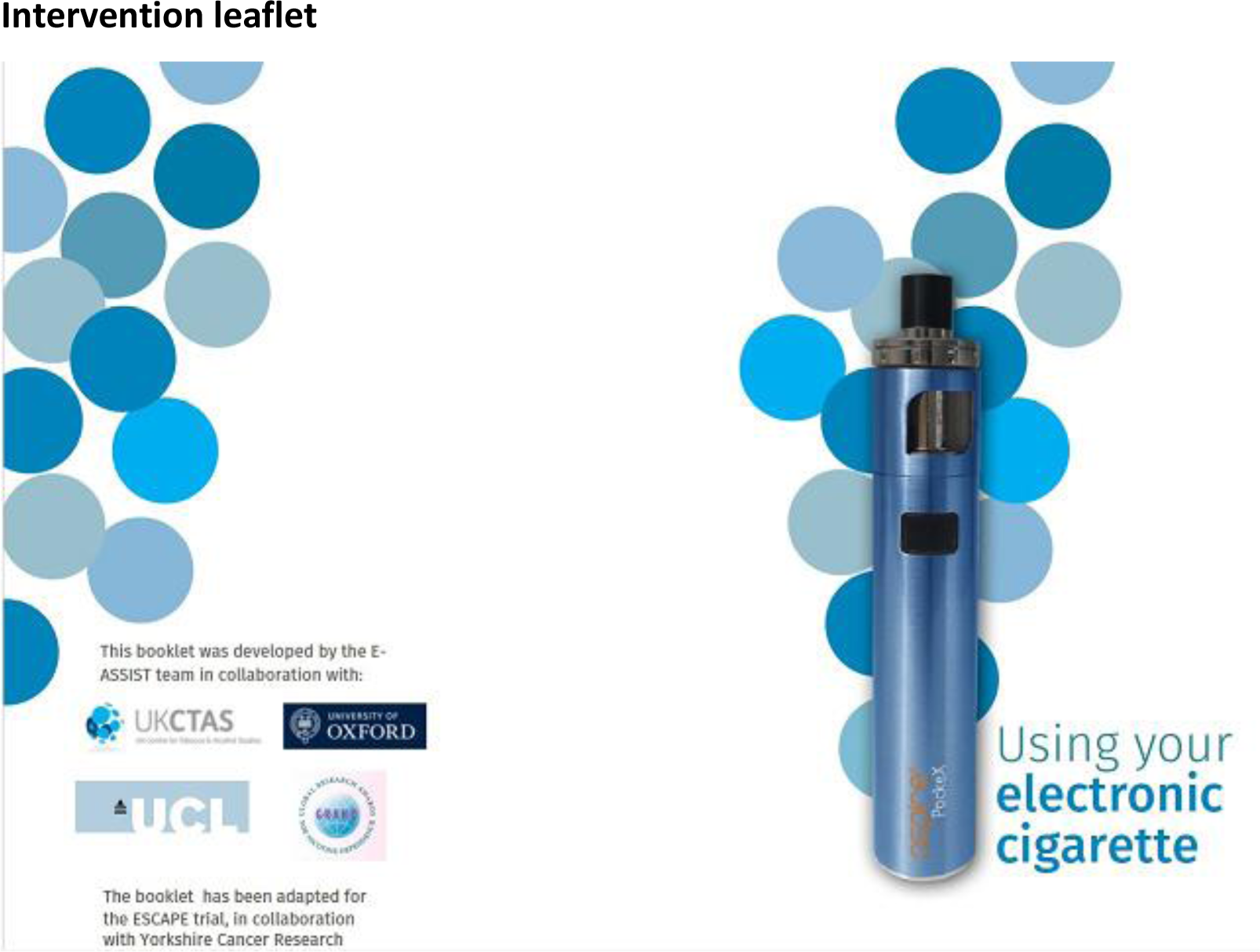

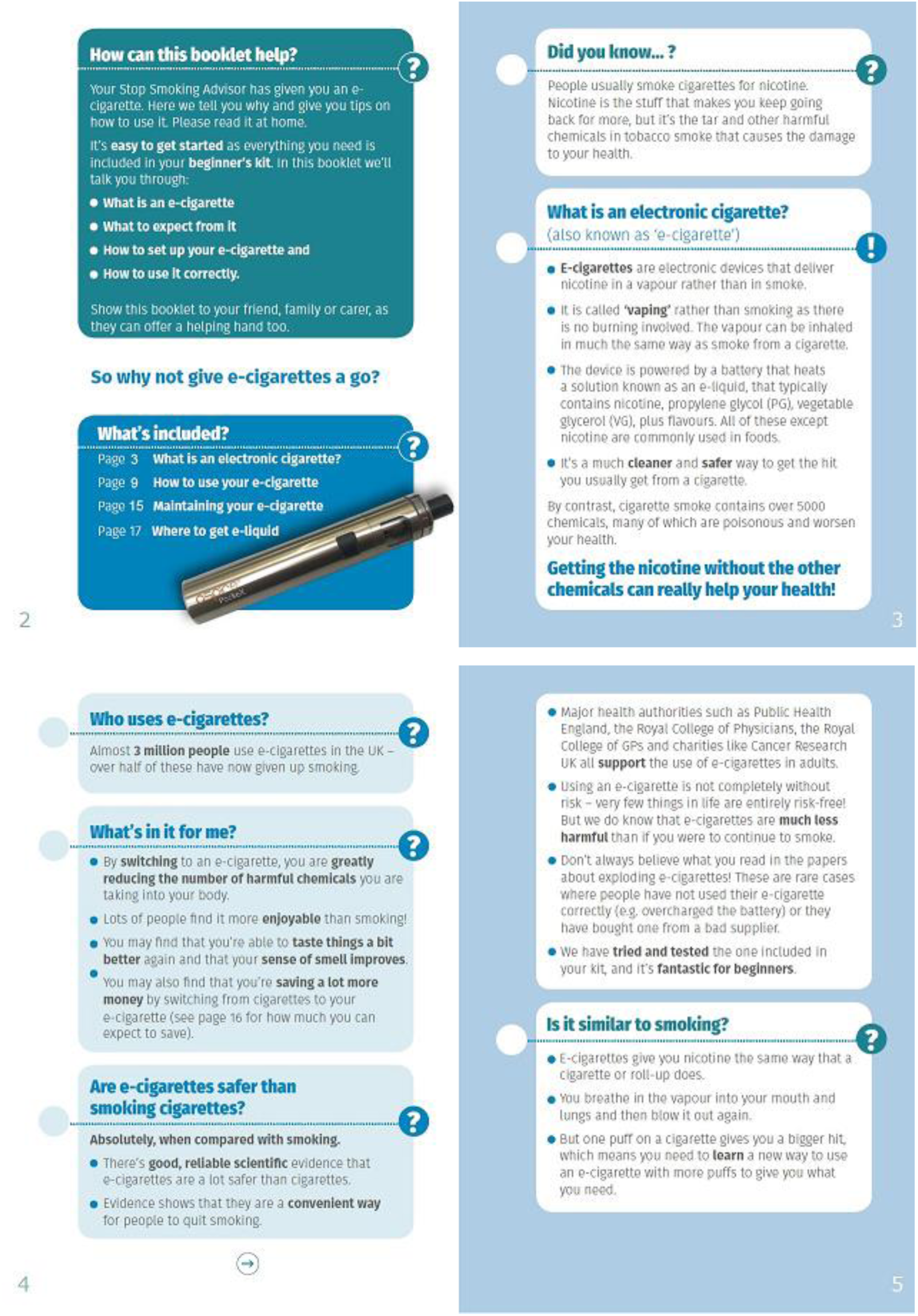

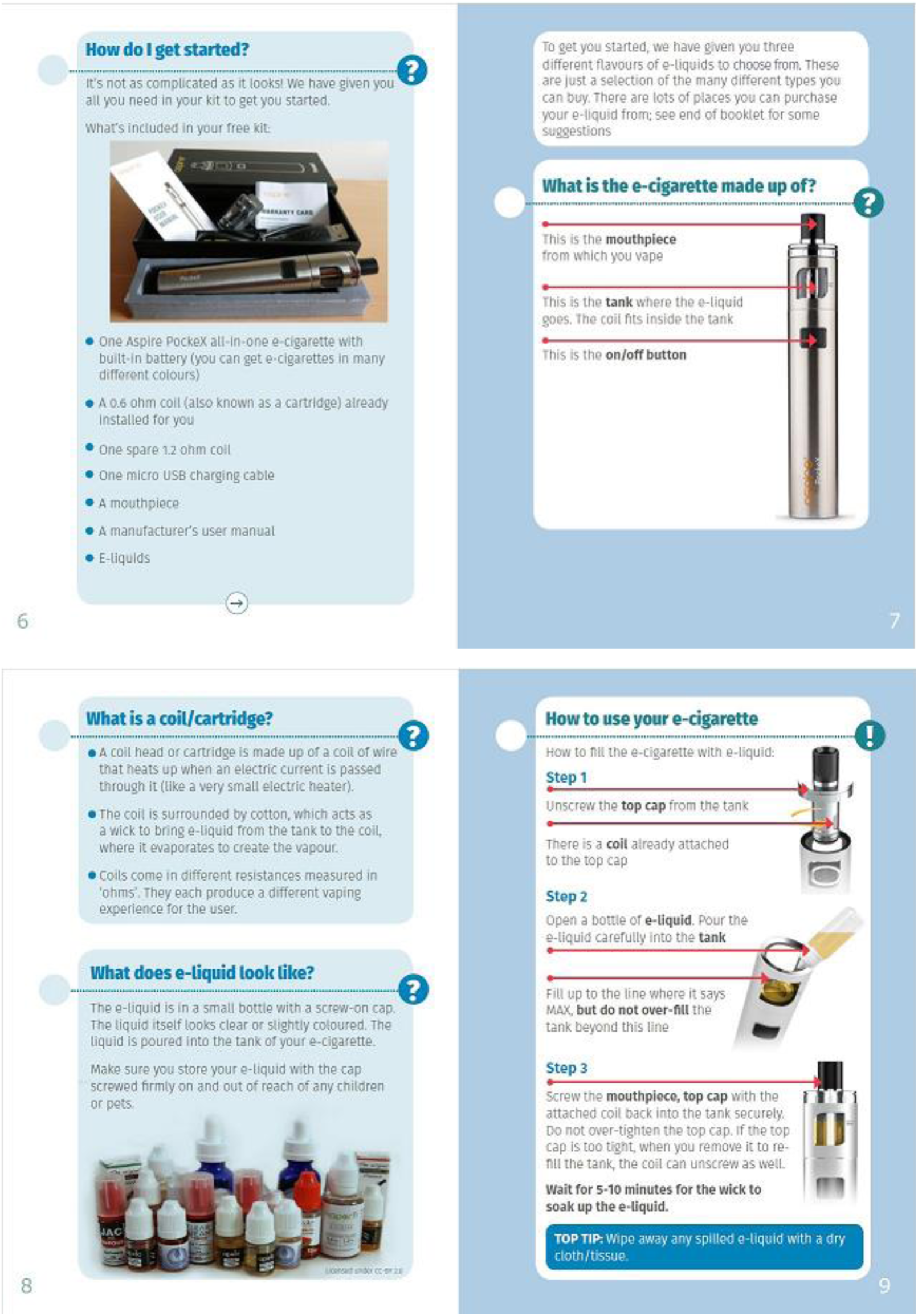

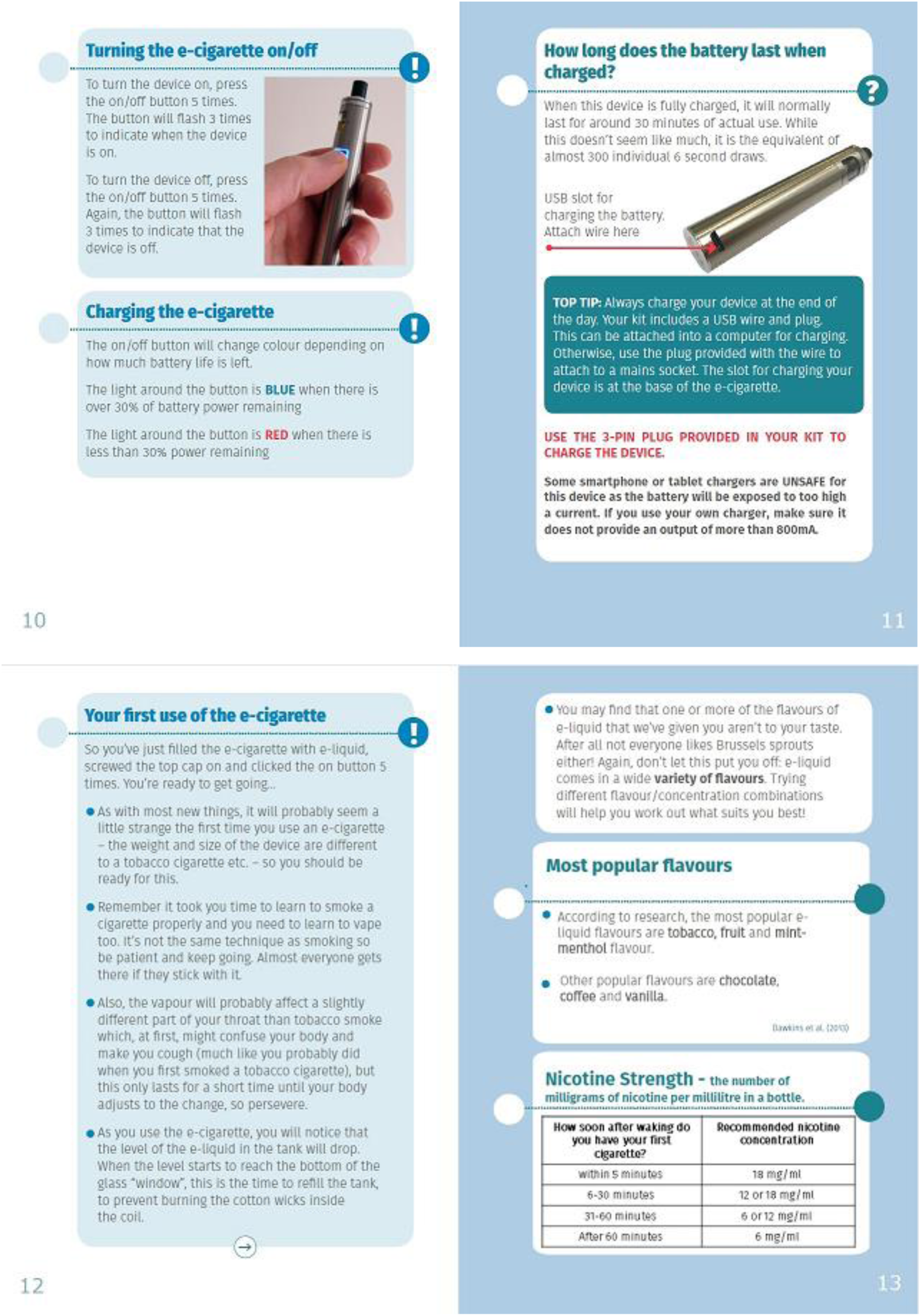

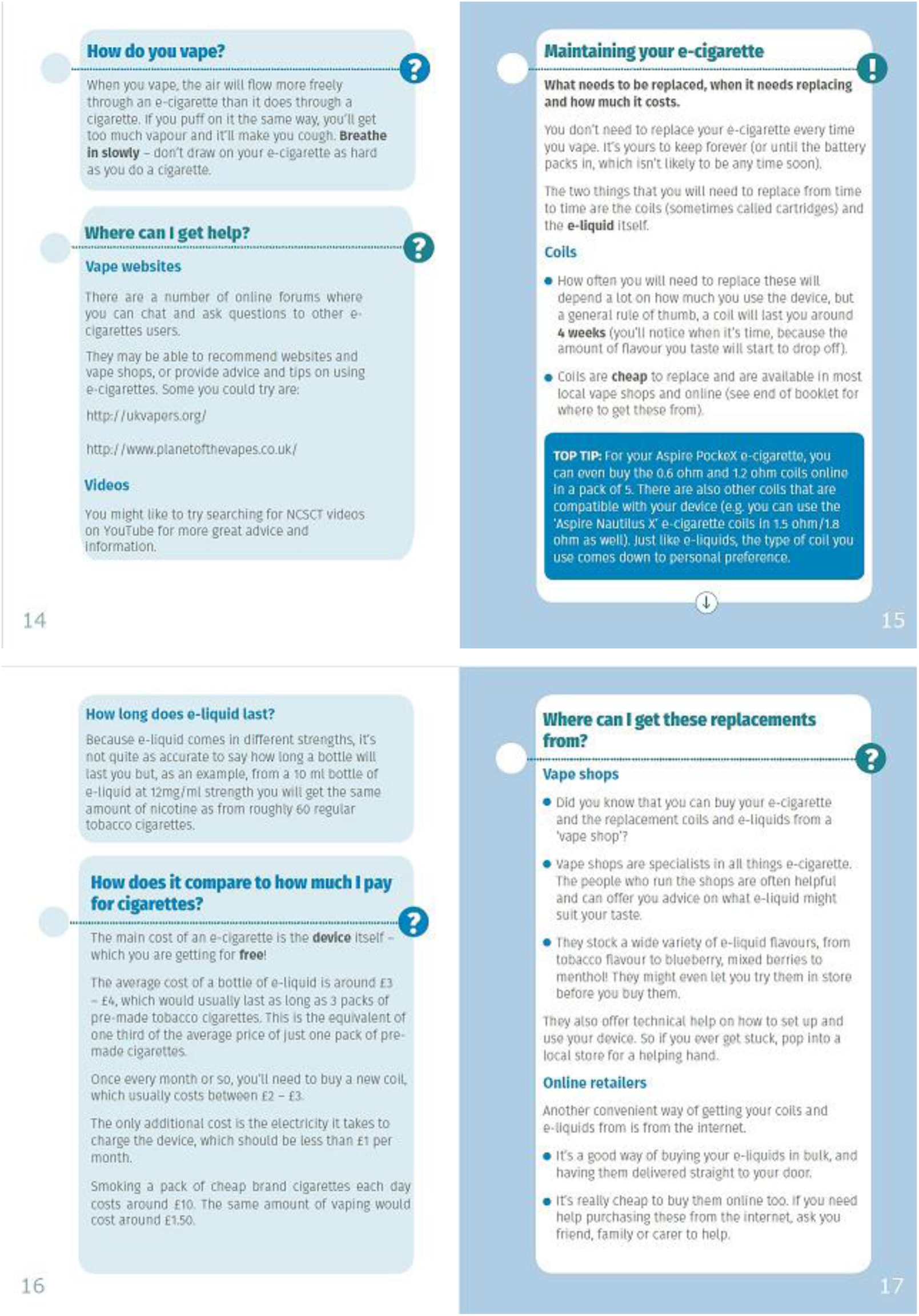

